# Investigating associations between physical multimorbidity clusters and subsequent depression: cluster and survival analysis of UK Biobank data

**DOI:** 10.1101/2024.07.05.24310004

**Authors:** Lauren Nicole DeLong, Kelly Fleetwood, Regina Prigge, Paola Galdi, Bruce Guthrie, Jacques D. Fleuriot

**Affiliations:** Artificial Intelligence and its Applications Institute, School of Informatics, University of Edinburgh, Edinburgh, UK; Usher Institute, University of Edinburgh, Edinburgh, UK; Advanced Care Research Centre, Usher Institute, University of Edinburgh, Edinburgh, UK

## Abstract

**Background:** Multimorbidity, the co-occurrence of two or more conditions within an individual, is a growing challenge for health and care delivery as well as for research. Combinations of physical and mental health conditions are highlighted as particularly important. The aim of this study was to investigate associations between physical multimorbidity and subsequent depression.

**Methods and Findings:** We performed a clustering analysis upon physical morbidity data for UK Biobank participants aged 37-73 years at baseline data collection between 2006-2010. Of 502,353 participants, 142,005 had linked general practice data with at least one physical condition at baseline. Following stratification by sex (77,785 women; 64,220 men), we used four clustering methods (agglomerative hierarchical clustering, latent class analysis, *k*-medoids and *k*-modes) and selected the best-performing method based on clustering metrics. We used Fisher’s Exact test to determine significant over-/under-representation of conditions within each cluster. Amongst people with no prior depression, we used survival analysis to estimate associations between cluster-membership and time to subsequent depression diagnosis.

The *k*-modes models consistently performed best, and the over-/under-represented conditions in the resultant clusters reflected known associations. For example, clusters containing an overrepresentation of cardiometabolic conditions were amongst the largest clusters in the whole cohort (15.5% of participants, 19.7% of women, 24.2% of men). Cluster associations with depression varied from hazard ratio (HR) 1.29 (95% confidence interval (CI) 0.85-1.98) to HR 2.67 (95% CI 2.24-3.17), but almost all clusters showed a higher association with depression than those without physical conditions.

**Conclusions:** We found that certain groups of physical multimorbidity may be associated with a higher risk of subsequent depression. However, our findings invite further investigation into other factors, like social ones, which may link physical multimorbidity with depression.

## Introduction

Multimorbidity, the simultaneous occurrence of two or more long-term conditions in an individual is increasingly common as populations age, and it challenges existing health systems ^1,2^. Multimorbidity is more common with increasing age, in women and in the less affluent ^3,4^. Studying the co-occurrence of multiple long-term conditions in the same individual has the potential to inform understanding of disease causation and support planning of current and future health and care services ^5–7^.

Depression affects millions of people worldwide ^8,9^ and is ranked by the World Health Organization as one of the most burdensome diseases ^9,10^. There is strong evidence that depression co-occurs with other mental health disorders ^11,12^, and several ongoing studies aim to identify potential shared mechanisms ^11,13^. However, previous studies have also found depression to be more common in people with particular chronic physical illnesses, such as cardiovascular disease ^14^, multiple sclerosis ^15^, and inflammatory bowel disease ^16^. Physical ill-health might cause depression because it creates psychological disturbance through ‘biographical disruption’ that threatens a sense of identity, or because of impact on physical or social function. Alternatively, physical conditions may cause depression through intermediate biological processes, like inflammation ^15,17^, in which case we might expect that different combinations or patterns of physical conditions would be more strongly associated with depression than others.

Several studies have used cluster analyses to identify common patterns of physical conditions ^18–21^, typically using one method, such as agglomerative hierarchical clustering ^18,19,22^, *k*-medoids ^20,23^, Latent Class Analysis ^21,24–26^, or *k*-means approaches ^27–30^. Additionally, since morbidity data is binary (a person has a condition or does not), some common clustering methods are inappropriate since they use similarity measures incompatible with categorical data ^18,31^. Therefore, the aim of this study was to explore and compare the use of four independent clustering methods appropriate for binary data and to examine whether certain groups of physical conditions are associated with the subsequent diagnosis of depression.

## Materials and methods

### Data selection and pre-processing

We used data from UK Biobank ^32^. Participants aged 37-73 years attended a baseline assessment during 2006-2010 which collected data on demography, lifestyle habits, health conditions, and a range of physical and laboratory measurements. Participants provided written informed consent for linkage to national datasets including general practice (GP) (primary care), hospital, cancer registry and death records ^32^. The UK Biobank has ethical approval from the NHS North West Research Ethics Committee (reference: 21/NW/0157).

To robustly ascertain a broad range of long-term conditions, our study population included participants with a continuous GP record from at least a year before to at least one day beyond their baseline assessment. We excluded records from the UKB extract of the Vision practice management system in England because the extraction process excluded participants who died prior to data extraction. We also excluded participants who withdrew from the study (Supplementary Fig. 1).

We ascertained the presence of depression and 69 long-term physical health conditions at baseline using data from the baseline visit and from linked GP, hospital, and cancer registry records based on previously published lists ^33,34^ (Supplementary Table 1). The UK National Health Service limits registration to one practice at a time, and GP records transfer between practices so should capture an individual’s entire medical history. However, available hospital and cancer registry records began at different times for England, Wales and Scotland. Therefore, we used all GP records up to baseline assessment date, and to be consistent across countries, we used hospital and cancer registry records within eight years before baseline assessment date. We used published codelists to identify diagnoses from GP records using Read V2 and CTV3 diagnosis codes, hospital records using ICD-10 diagnosis codes and OPCS-4 procedure codes, and cancer registry records using ICD-10 codes ^34^. We similarly ascertained depression during follow-up using information from GP, hospital and death records. Eligible participants with no history of depression prior to baseline were followed up to the earliest of depression diagnosis, death or the end of their available GP or hospital records.

### Models and metrics

We explored the suitability of four methods (*k*-modes ^35,36^, *k-*medoids ^23^, Latent Class Analysis (LCA) ^24^, and agglomerative hierarchical clustering (AHC) ^22^ (Supplementary Appendix 1)) to cluster all participants based on binary features denoting the absence/presence of the 69 baseline physical conditions. Participants with no physical conditions at baseline were excluded from the clustering analysis. We additionally clustered separately for men (all 69 conditions) and women (67 conditions since erectile dysfunction and hyperplasia of the prostate are only found in men), as well as in the whole population, because of known sex differences in patterns of individual morbidities and multimorbidity ^37,38^.

To select the number of clusters for each method, we used various heuristics, including the elbow method on a scree plot ^39^ for both *k*-modes and *k*-medoids, the minimal Bayesian Information Criterion ^40^ for LCA, and Hamming distance ^41^ for AHC. To assess suitability and performance of these clustering methods, we used three performance metrics (Calinski and Harabasz score ^42^, Davies-Bouldin score ^43^, and Silhouette score ^44^ (Supplementary Appendix 1)), which are appropriate for unsupervised clustering. Since *k*-modes and LCA are sensitive to differences in initialization ^24,45^, we repeated them five times and compared with other models using the mean and standard deviation across the five experiments.

Thereafter, we used two metrics to analyze over-and under-representation of conditions in each cluster. We designed one metric, the *adjusted relative frequency (ARF)* to measure the magnitude of over-or under-represented conditions within a cluster, relative to prevalence in the whole cohort. For each condition, ARF is calculated as:

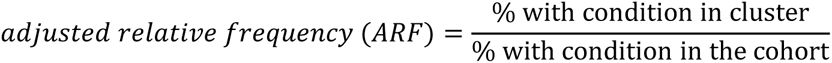

An ARF of exactly one indicates that the condition occurs at the same relative frequency as it does in the entire cohort, and values greater or less than one indicate over-and under-representation, respectively. We used Fisher’s Exact Test (two-sided, α=0.05) to evaluate whether over-or under-representation of a condition in each cluster was statistically significant, using a Bonferroni correction to account for multiple testing ^46–48^. Finally, we visualised and compared statistically significant results on a *bubble heatmap* (Supplementary Appendix 2). To allow others to conduct similar cluster analyses, we made the code available as a software package (https://github.com/laurendelong21/clusterMed).

### Survival analysis to predict depression diagnosis

Using participants without a record of depression at baseline, we applied Cox regression models ^49^ to evaluate time to depression diagnosis by condition cluster, accounting for death as a competing risk ^50^. Participants with no physical conditions at baseline were included as the reference group. We ran separate models for the whole cohort and for men and women separately, examining associations between cluster membership and subsequent depression. All models were adjusted for baseline age, ethnicity, country of residence and deprivation. The model for the whole cohort was additionally adjusted for sex. Baseline age was included in the models as a continuous variable; all other variables were categorical. Ethnicity was self-reported at baseline, and we categorized it into five groups (Black, Mixed, South Asian, White, and any other ethnic group ^51^). Country of residence (England, Wales or Scotland) and area-based deprivation, measured by the Townsend Deprivation Index ^52^, were derived from participants’ home addresses at baseline. We divided the Townsend Deprivation Index into deciles within the entire UK Biobank cohort. A small number of participants (368 women and 435 men) who were missing data on ethnicity, country or deprivation were excluded from the survival analyses.

## Results

### Performance metrics across various clustering methods

There were 140,956 participants (73,036 women and 67,920 men) with at least one physical condition at baseline who were included in the clustering analysis (Supplementary Fig. 1). Performance metrics for each of the four methods explored are reported in Table 1.

**Table 1.** Performance metrics, number of clusters, and cluster sizes across four clustering methods.

| Cohort | Clustering method | Calinski and Harabasz score (higher is better) | Davies-Bouldin score (lower is better) | Silhouette score (higher is better) | No. of clusters | No. of singleton clusters | Median participants / cluster | (Min-max.) participants per cluster |
| --- | --- | --- | --- | --- | --- | --- | --- | --- |
| Whole | LCA | 5567.79 ± 175.06 | 3.80 ± 0.34 | 0.08 ± 0.03 | 6 | 0 | 17203.5 | (6816 - 57564) |
|  | AHC | 628.39 | 4.38 | 0.09 | 10 | 0 | 2495.5 | (11 - 110366) |
|  | *k*-medoids | 1270.00 | 2.48 | 0.08 | 25 | 17 | 1 | (1 - 55553) |
|  | *k*-modes | 6079.53 ± 1228.07 | 2.43 ± 0.16 | 0.17 ± 0.02 | 8 | 0 | 14954.5 | (974 - 39601) |
| Women-only | LCA | 4098.56 ± 5.23 | 3.53 ± 0.00 | 0.15 ± 0.00 | 5 | 0 | 15794 | (2373 - 30529) |
|  | AHC | 524.62 | 4.82 | 0.04 | 10 | 0 | 1374.5 | (2 - 47728) |
|  | *k*-medoids | 3672.43 | 2.60 | 0.14 | 6 | 0 | 9472 | (3942 - 28757) |
|  | *k*-modes | 3456.82 ± 555.59 | 2.36 ± 0.10 | 0.16 ± 0.04 | 8 | 0 | 9233.5 | (562 - 22787) |
| Men-only | LCA | 3134.68 ± 91.36 | 3.52 ± 0.02 | 0.10 ± 0.00 | 5 | 0 | 13868 | (6827 - 16448) |
|  | AHC | 274.95 | 4.18 | 0.20 | 10 | 0 | 1709 | (7 - 27353) |
|  | *k*-medoids | 1225.80 | 2.17 | 0.11 | 13 | 7 | 1 | (1 - 25595) |
|  | *k*-modes | 2779.42 ± 463.66 | 2.35 ± 0.07 | 0.15 ± 0.03 | 8 | 0 | 8545.5 | (649 - 15532) |

157 Results for LCA and *k*-modes are reported as average ± standard deviation across five independent models.

Models based on AHC consistently achieved the poorest Calinski and Harabasz, and Davies-Bouldin scores in all three cohorts. Models based on LCA had better metrics than AHC-based models, with particularly high Calinski and Harabasz scores, but the Davies-Bouldin scores were consistently worse in comparison to *k-*modes or *k-*medoids based models. In contrast, the best Davies-Bouldin scores were achieved by the *k-*modes and *k-*medoids based models. However, since the Davies-Bouldin score assesses similarity between the most similar clusters, scores may be optimistic when several clusters only contain a single (or very few) participant(s). This was the case for the *k*-medoids models for the whole and men-only cohorts. The presence of singleton clusters is also concurrent with a larger number of total clusters. Specifically, all three models based on *k*-modes discovered eight clusters, all models using AHC discovered ten clusters, and all models using LCA discovered five or six clusters. In contrast, the *k-* medoids models discovered 25 clusters for the whole population (17 only had one participant, six for women-only (no singletons), and 13 for men-only (seven singletons) (Table 1). Therefore, while *k-* medoids models had comparable Davies-Bouldin scores to *k-*modes models, the results were less informative and consistent. For each cohort, we therefore selected the best performing *k*-modes model with the highest Calinski and Harabasz score among the five independent runs.

### Differential representation of physical conditions within *k*-modes clusters

Many of the significantly over-represented conditions within several clusters aligned with body systems (Fig. 1) and we therefore used clinical judgement to name the clusters according to the systems or conditions which were most prominent (Supplementary Tables 2-5). The four largest clusters in whole cohort are *Mixed including cancer* (27.9% of participants in the cohort), *Healthy + Rhinitis* (22.2%), *Cardiovascular disease (CVD) + diabetes* (15.5%), and *Very extensive morbidity* (12.5%). For women, the four largest clusters are *Mixed including cancer* (29.3%), *CVD + diabetes* (19.7%), *Musculoskeletal (MSK)* (16.4%), and *Healthy + Rhinitis* (15.9%). Finally, for men, the four largest clusters are *CVD + diabetes* (24.2%), *Mixed including cancer* (20.8%), *MSK + others* (19.1%), and *Healthy + Rhinitis* (17.2%) (Fig. 2, Table 2).

**Fig. 1.**
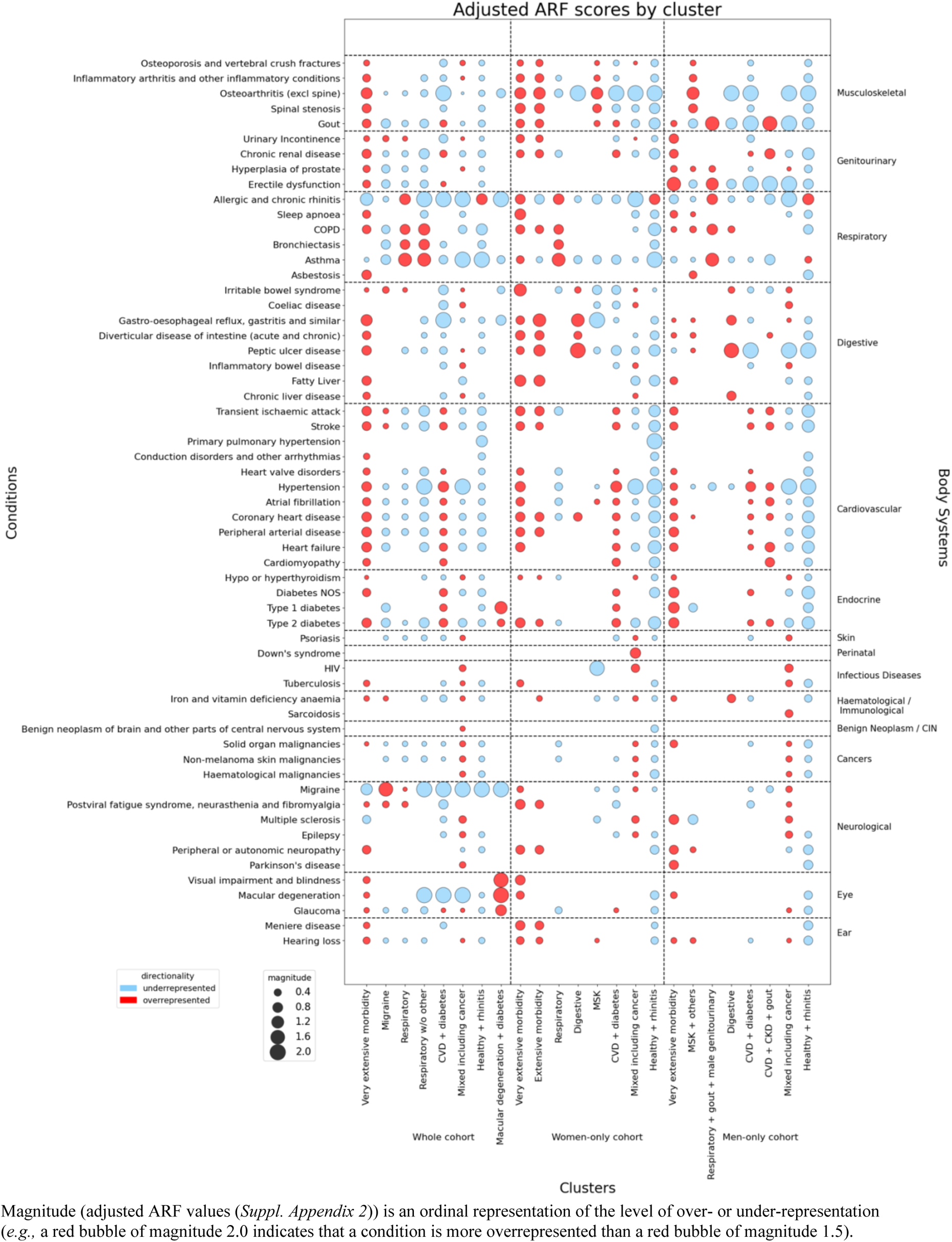
Bubble heatmap shows under-and over-represented conditions in each cluster from the k-modes derived models.

**Fig. 2.**
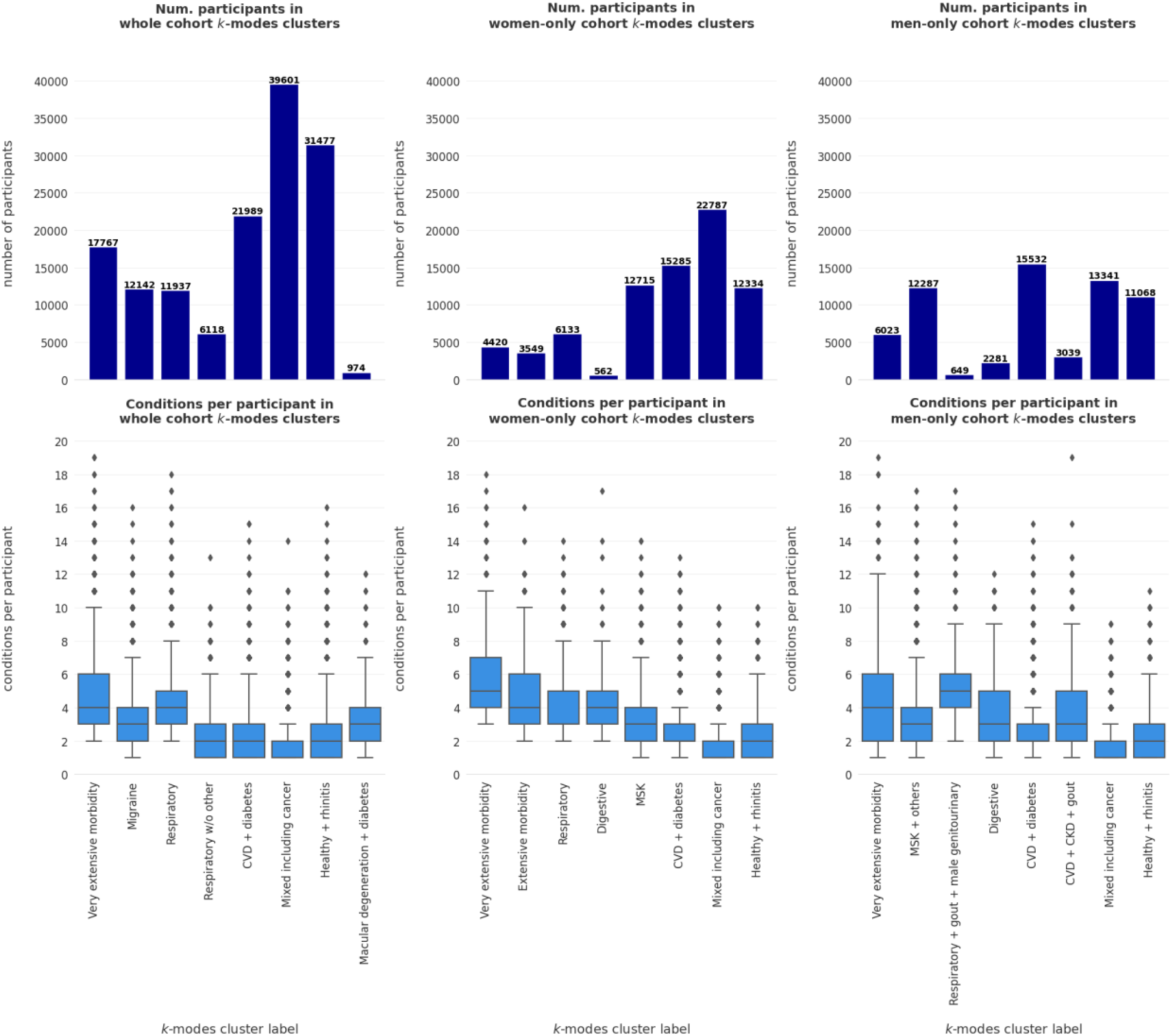
Cluster sizes and condition counts.

**Table 2.** Hazard ratios per cluster for the development of subsequent depression.

| **Cohort** | **Cluster Label** | **No. (%) of participants in cluster** | **Mean no. of conditions per participant in cluster** | **Hazard ratio (95% CI)** |
| --- | --- | --- | --- | --- |
| Whole cohort | Very extensive morbidity | 17767 (12.5) | 4.50 | 2.42 (2.17, 2.69) |
|  | Migraine | 12142 (8.6) | 3.23 | 1.96 (1.75, 2.19) |
|  | Respiratory | 11937 (8.4) | 4.00 | 1.95 (1.74, 2.18) |
|  | Respiratory w/o other | 6118 (4.3) | 2.29 | 1.86 (1.62, 2.15) |
|  | CVD + diabetes | 21989 (15.5) | 2.38 | 1.78 (1.61, 1.97) |
|  | Mixed including cancer | 39601 (27.9) | 1.77 | 1.62 (1.48, 1.77) |
|  | Healthy + rhinitis | 31477 (22.2) | 2.58 | 1.59 (1.46, 1.75) |
|  | Macular degeneration + diabetes | 974 (0.7) | 3.09 | 1.29 (0.85, 1.98) |
| Women-only | Very extensive morbidity | 4420 (5.7) | 5.69 | 2.67 (2.24, 3.17) |
|  | Extensive morbidity | 3549 (4.6) | 4.61 | 2.56 (2.12, 3.10) |
|  | Respiratory | 6133 (7.9) | 3.72 | 1.93 (1.67, 2.23) |
|  | Digestive | 562 (0.7) | 4.49 | 1.83 (1.14, 2.93) |
|  | MSK | 12715 (16.4) | 2.85 | 1.81 (1.58, 2.07) |
|  | CVD + diabetes | 15285 (19.7) | 2.70 | 1.71 (1.52, 1.94) |
|  | Mixed including cancer | 22787 (29.3) | 1.75 | 1.63 (1.46, 1.82) |
|  | Healthy + rhinitis | 12334 (15.9) | 2.02 | 1.48 (1.30, 1.67) |
| Men-only | Very extensive morbidity | 6023 (9.4) | 4.30 | 2.65 (2.22, 3.18) |
|  | MSK + others | 12287 (19.1) | 3.46 | 2.50 (2.15, 2.91) |
|  | Respiratory + gout + male genitourinary | 649 (1.0) | 5.23 | 2.25 (1.43, 3.53) |
|  | Digestive | 2281 (3.6) | 3.47 | 2.06 (1.60, 2.66) |
|  | CVD + diabetes | 15532 (24.2) | 2.71 | 1.90 (1.65, 2.20) |
|  | CVD + CKD + gout | 3039 (4.7) | 3.77 | 1.87 (1.47, 2.38) |
|  | Mixed including cancer | 13341 (20.8) | 1.62 | 1.60 (1.38, 1.86) |
|  | Healthy + rhinitis | 11068 (17.2) | 2.08 | 1.50 (1.29, 1.75) |

All models were adjusted for baseline age, ethnicity, country of residence, and deprivation.

Of the remaining clusters, there were some similarities across all three cohorts (*e.g. Respiratory* clusters). Generally, clusters with more participants also tended to have fewer conditions per participant (Fig. 2). For example, the *Mixed including cancer* clusters had the lowest mean conditions per participant (whole: 1.77; women: 1.75; men: 1.62). Such clusters may serve as "miscellaneous" categories for participants with condition profiles that are not easily grouped and/or people with one dominant condition. However, there were also differences. For example, there were clusters which only appeared in the whole population (*Migraine*) and clusters which only appeared in the sex-stratified cohorts (*Digestive* and *MSK* clusters).

### Subsequent incident depression per identified cluster

Analysis of time to incident depression diagnosis included 141,001 participants (73,036 women and 67,920 men), excluding 30,770 participants with a history of depression at baseline (20,592 women and 10,178 men). In addition to participants included in the clustering analysis, this analysis also included 30,551 participants with no physical conditions at baseline (16,238 women and 14,313 men) (Supplementary Fig. 1). During an average follow-up of 6.8 years, 5,904 (4.2%) participants, including 3,574 (4.9%) women and 2,330 (3.4%) men, had a new depression diagnosis. Generally, participants with physical conditions at baseline had a higher rate of subsequent depression than participants with no physical conditions at baseline (Table 2, Supplementary Fig. 3).

There were several consistencies across cohorts. The *Very extensive morbidity* clusters were the most strongly associated with depression in all three cohorts (whole: HR 2.42, 95% CI 2.17-2.69; women: HR 2.67, 95% CI 2.24-3.17; men: HR 2.65, 95% CI 2.22-3.18). Additionally, the *Healthy + rhinitis* (whole: HR 1.59, 95% CI 1.46-1.75; women: HR 1.48, 95% CI 1.30-1.67; men: HR 1.50, 95% CI 1.29-1.75) and *Mixed including cancer* (whole: HR 1.62, 95% CI 1.48-1.77; women: HR 1.63, 95% CI 1.46-1.82; men: HR 1.60, 95% CI 1.38-1.86) clusters were generally the most weakly associated. Finally, association with depression also appeared to increase with the number of conditions per participant (Fig. 3). However, there were some exceptions; for example, the whole cohort’s *Macular degeneration + diabetes* cluster had the fourth highest mean number of conditions (3.09), but was only weakly associated with depression (HR 1.29, 95% CI 0.85-1.98).

**Fig. 3.**
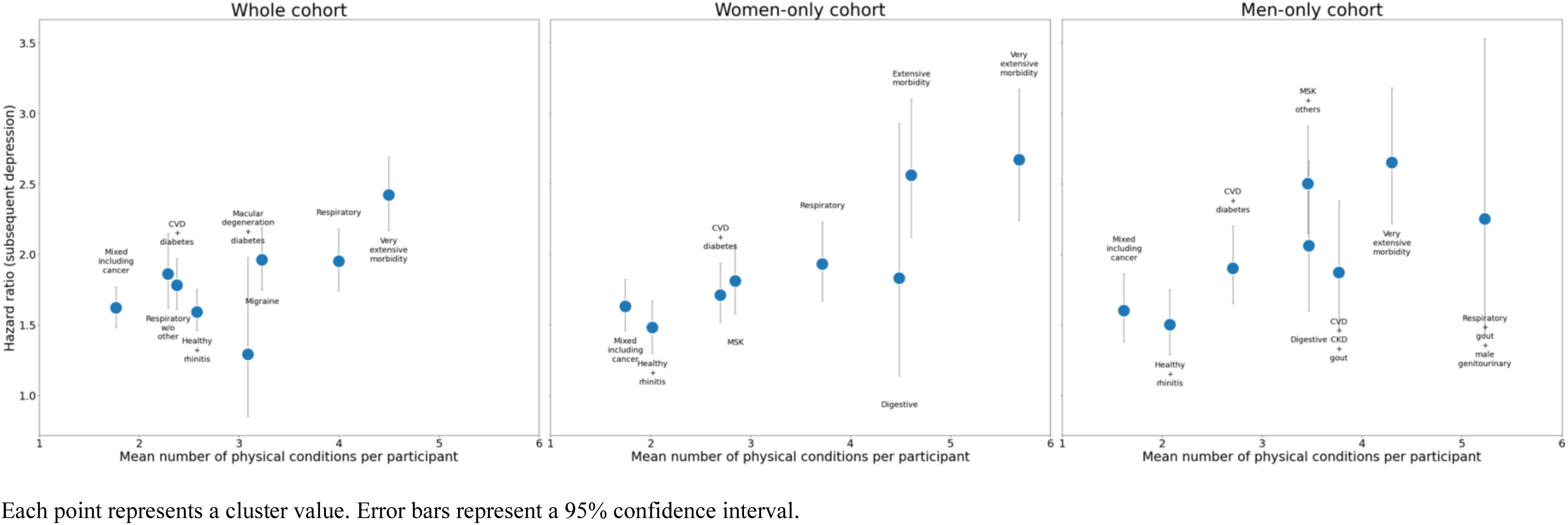
Risk of subsequent depression by mean number of physical conditions.

## Discussion

### Summary of findings

This study systematically explored clustering of physical health conditions using four methods appropriate for binary data (*k*-modes ^35,36^, *k-*medoids ^23^, Latent Class Analysis ^24^, and agglomerative hierarchical clustering (AHC) ^22^). *K*-modes performed best, and the clusters identified were reasonably interpretable and often aligned with known associations between conditions. People with any physical condition at baseline were generally more likely to develop depression than people without any physical condition. There was some variation in this association by cluster which may be at least partly driven by differences in the mean number of physical conditions in each cluster.

### Comparison with other studies

Existing studies of morbidity clustering typically apply a single method. One study compared LCA to a Bayesian, network-based approach, but used age and admission type, rather than conditions alone, to drive cluster formation ^53^. Two other studies explored AHC and *k*-means in the same dataset, but chose *k*-means on the basis of AHC being too computationally intensive rather than based on performance ^27,28^. Additionally, despite the use of *k*-means ^54^ by several multimorbidity studies ^27–30^, it typically relies upon Euclidean distance as its similarity measure ^31^, which is unsuitable for binary data ^18^. Other multimorbidity studies have used *k*-means *after* a Multiple Correspondence Analysis ^55,56^, which represents categorical features as a low-dimensional Euclidean space ^29,30^. While this transforms the data features into an appropriate format for *k*-means, it also manipulates the data based on their pairwise co-occurrences, which may not be appropriate for every dataset.

This study finds that almost all physical morbidity clusters are associated with higher risk of subsequent depression than the group with no physical conditions at baseline. Although the strength of association varied by cluster, this seemed to be partly explained by the mean number of conditions in the cluster. This is consistent with a similar study ^29^ which found associations between severe mental illness and a higher number of physical conditions. Another similar study, which aimed to identify groups of physical conditions associated with incident depression within a Taiwanese cohort, also found that social factors played a role on the risk of subsequent depression diagnosis ^21^. Specifically, they found that amongst four *Cardiometabolic*, *Arthritis-cataract*, *Multimorbidity*, and *Relatively healthy* clusters, those within the *Arthritis-cataract* and *Multimorbidity* clusters had significantly higher risk of depression than healthy individuals. However, this association was attenuated for participants who engaged in social activities, including a job, volunteer experience, or community activities ^21^.

### Strengths and limitations

Strengths of this study include the analysis of a large dataset which records morbidities in both baseline research data and linked routine data, as well as the inclusion of a wide set of morbidities recommended by a recent consensus study ^57^. Notably, this study is unique for its implementation and comparison between four clustering methods appropriate for binary data. A limitation is that the data are collected from volunteers who are generally more affluent than the UK average, and people from ethnic minorities are somewhat under-represented ^58^. Additionally, there is no standard way to evaluate the validity of identified clusters, although the observed clusters do include several known clinical associations. Consequently, this warrants further validation studies in other datasets to explore reproducibility of cluster solutions.

### Implications for research

Many previous studies of morbidity clustering do not provide much information about which conditions are over or under-represented in clusters, which leaves readers relying solely on author-chosen cluster labels for interpretation ^19,21^. For example, it is common for other studies to identify a ‘cardiometabolic’ cluster ^21^ and the *CVD + diabetes* cluster in our study was amongst the three largest clusters in all three cohorts. However, it is not straightforward to compare clusters across studies because of considerable variation in the conditions included in analysis, and because many clustering studies do not provide detailed information about the nature of identified clusters. Key implications are that clustering studies should be more consistent in the choice of conditions to include (and, at a minimum, follow consensus recommendations ^57^). Additionally, they should report the nature of clusters to help understand them beyond their high-level labels (by, for example, visualizing the prevalence of individual conditions in each cluster alongside over/under representation). We believe that our Adjusted Relative Frequency (ARF) measure with visualization in a bubble heatmap demonstrates one way to do this. However, there is a need for multimorbidity researchers to develop improved and consistent cluster visualizations and explanations to facilitate interpretation and to enhance clinical utility.

Morbidity clustering studies also typically use one clustering method, but there is no single clustering method which is likely to be optimal for every dataset. We therefore believe that clustering studies should more systematically explore different methods and make explicit how they choose the best method for their datasets and purposes. To encourage similar systematic comparison of different cluster methods, we have provided access to our code (https://github.com/laurendelong21/clusterMed). Many studies also cluster the entire population which is likely not sensible given the very different incidence and prevalence of disease with age and, to a lesser extent, sex and ethnicity. In this analysis, study participants were mostly middle-aged (so we did not further stratify by age) and overwhelmingly white, but we found some differences in the clusters identified in the whole population versus women or men separately. Although whole population clustering may be appropriate in some circumstances, reporting of clusters stratified by age and sex (and ethnicity if the data permits) would be valuable to explore how clustering varies by demographic characteristics.

Finally, further research to better understand why physical multimorbidity is associated with subsequent depression is needed. The general trend between increased risk of subsequent depression and mean number of conditions suggests a social explanation: suffering more conditions may more strongly interrupt one’s life or sense of self. However, the relationship between depression and physical conditions is very likely bidirectional and longitudinal research which better examines how the two interact over a lifetime would be valuable.

## Conclusions

Using the best performing of four different clustering methods, this study identified several multimorbidity clusters which align with known clinical associations. Association with depression varied between clusters, but this may be partly driven by differences in the number of conditions. More research is needed to better understand the mechanisms underlying such associations.

## Data Availability

The UK Biobank data is not openly available to protect the rights of participants. Researchers can register for access here: https://www.ukbiobank.ac.uk/enable-your-research/register.

## Code Availability

Corresponding code is available at https://github.com/laurendelong21/clusterMed.

## Author Contributions

All authors contributed to writing the manuscript. L.N.D. and K.F. conducted the analyses and made the figures. L.N.D. and P.G. wrote the software. K.F. and R.P. processed and prepared the data. J.D.F. and

B.G. designed and supervised the study.

## Data Availability

The UK Biobank data is not openly available to protect the rights of participants. Researchers can register for access here: http://www.ukbiobank.ac.uk/enable-your-research/register.

## Acknowledgments

This work was co-funded by the Medical Research Council and the National Institute for Health Research (grant number MC/S028013), and the NIHR AIM-CISC programme (grant number NIHR202639). The views expressed are those of the author(s) and not necessarily those of the NIHR or the Department of Health and Social Care. The study was conducted using the UK Biobank Resource under application number 57213.

LND is individually funded by a Global Informatics Scholarship from the School of Informatics at the University of Edinburgh. The School of Informatics had no role in study design, data collection and analysis, decision to publish, or preparation of the manuscript.

The authors would like to thank the UK Biobank participants and the UK Biobank staff for their contributions to this study. The authors would like to thank the public members of our advisory board, Dr Paul Kelly and Pat Watson, for providing thoughtful feedback throughout our project. This work has made use of the resources provided by the Edinburgh Compute and Data Facility (ECDF) (http://www.ecdf.ed.ac.uk/).

## Supporting information

**Supplementary Table 1.** 69 physical conditions with corresponding bodily systems.

| System | Condition |
| --- | --- |
| Skin conditions | Psoriasis |
| Perinatal conditions | Down's syndrome |
| Neurological conditions | Migraine<br>Peripheral or autonomic neuropathy<br>Epilepsy<br>Cerebral Palsy<br>Motor neuron disease<br>Multiple sclerosis<br>Myasthenia gravis<br>Parkinson's disease<br>Postviral fatigue syndrome, neurasthenia and fibromyalgia |
| Musculoskeletal conditions | Inflammatory arthritis and other inflammatory conditions<br>Gout<br>Osteoporosis and vertebral crush fractures<br>Osteoarthritis (excl spine)<br>Spinal stenosis |
| Mental Health Disorders | Dementia |
| Infectious Diseases<br>Infectious Diseases | Tuberculosis<br>HIV |
| Haematological/Immunological conditions | Sarcoidosis<br>Iron and vitamin deficiency anaemia<br>Immunodeficiencies<br>Sickle-cell anaemia<br>Thalassaemia |
| Diseases of the Respiratory System | Sleep apnoea<br>COPD<br>Bronchiectasis<br>Asthma<br>Asbestosis<br>Allergic and chronic rhinitis |
| Diseases of the Genitourinary system | Chronic renal disease<br>Urinary Incontinence<br>Erectile dysfunction<br>Non-acute cystitis<br>Hyperplasia of prostate |
| Diseases of the Eye | Visual impairment and blindness<br>Macular degeneration<br>Glaucoma |
| Diseases of the Endocrine System | Hypo or hyperthyroidism<br>Addisons disease<br>Cystic Fibrosis<br>Diabetes NOS |
|  | Type 1 diabetes<br>Type 2 diabetes |
| Diseases of the Ear | Meniere disease<br>Hearing loss |
| Diseases of the Digestive System | Fatty Liver<br>Chronic liver disease<br>Peptic ulcer disease<br>Irritable bowel syndrome<br>Diverticular disease of intestine (acute and chronic)<br>Gastro-oesophageal reflux, gastritis and similar<br>Coeliac disease<br>Inflammatory bowel disease |
| Diseases of the Circulatory System | Conduction disorders and other arrhythmias<br>Coronary heart disease<br>Cardiomyopathy<br>Heart valve disorders<br>Atrial fibrillation<br>Transient ischaemic attack<br>Peripheral arterial disease<br>Hypertension<br>Heart failure<br>Primary pulmonary hypertension<br>Stroke |
| Cancers | Solid organ malignancies<br>Haematological malignancies<br>Non-melanoma skin malignancies |
| Benign Neoplasm/CIN | Benign neoplasm of brain and other parts of central nervous system |

**Supplementary Figure 1.**
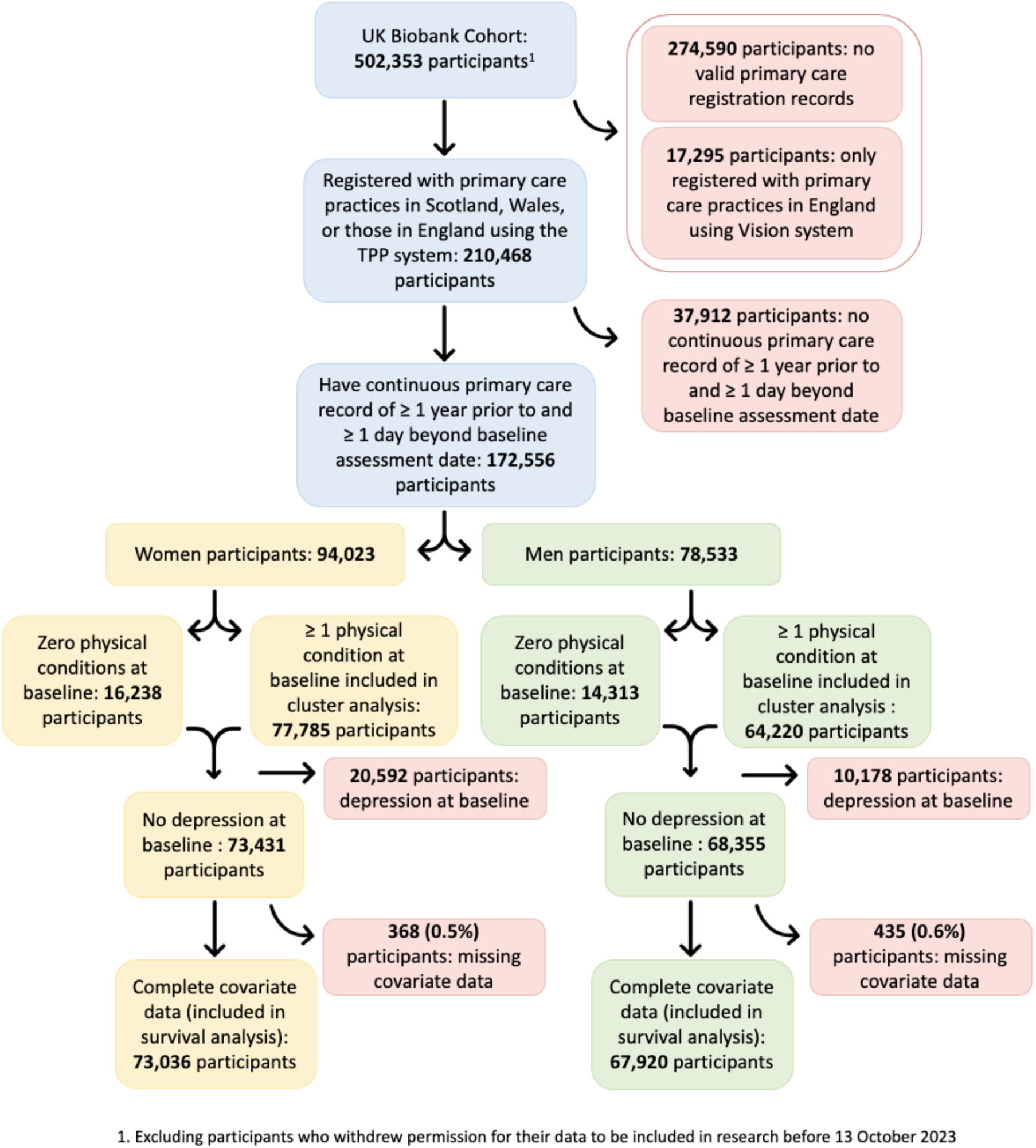
Flow diagram explaining data filtration steps.

**Supplementary Appendix 1.** Methodological Descriptions.

To identify a clustering method best suited for our data, we explored several combinations of metrics and methods which were, in theory, capable of handling the binary nature of the morbidity data. Specifically, we used the following metrics within this study:

- **Hamming distance** ^1^: This is a dissimilarity metric denoting the number of mismatching categories between two objects. It is formally defined as ^2^:

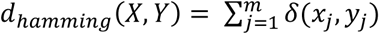

where:

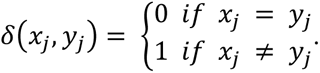
- **cosine similarity** ^3^: This is a measure which was originally used to indicate how similar two vector angles are to one another. For binary data, it can be understood as the number of true, or positive features are shared between two objects, divided by the product of the number of true objects from each object. Formally, it is defined as ^4^:

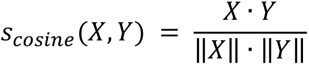

With these metrics, we explored the following four clustering methods:

- ***k*-modes** ^5,6^: This uses the same methodology as *k*-means clustering ^7^, but the centroid of each cluster is defined on the number of matching categories between data points, computed via the Hamming distance ^1^. In other words, the centroid represents the mode of the cluster, rather than the mean ^5,6^. Centroid initialization was performed via the frequency-based *Huang* metric ^5^.
- ***k*-medoids** ^8^: As above, this uses the same methodology as *k*-means clustering ^7^, but the centroid is an actual data point acting as the "median", computed via cosine similarity.
- **Latent Class Analysis (LCA)** ^9^: This aims to find groups or subtypes of cases (latent classes) in multivariate categorical data. It gives probabilities of class membership, rather than concrete class assignments, which are unique, so the user can see the likelihood that a data point truly belongs to its assigned class ^9^.
- **agglomerative hierarchical clustering (AHC)** ^10^: This is best understood as a "bottom-up" approach in which samples start out alone, then merge to form larger and larger clusters ^11^. We used a *complete* linkage (the maximum distance between points in two clusters ^12^), computed via Hamming distance ^1^.

Finally, we assessed cluster performance, including separation and overlap, with the following three performance metrics:

- **Calinski and Harabasz score** ^13^: This is the ratio of between-cluster dispersion to within-cluster dispersion. A higher Calinski and Harabasz score indicates better performance.
- **Davies Bouldin score** ^14^: This is a measure of cluster similarity to each cluster’s most similar cluster. A Davies Bouldin score closer to zero indicates better performance.
- **Silhouette score** ^15^: This is a measure of cluster fit which accounts for the mean distance between points in each individual cluster as well as the mean distance to points in the closest neighboring cluster. A silhouette score closer to one indicates better performance. Hamming distance was utilized as the distance metric.

The best similarity or dissimilarity metrics were selected for *k*-modes, *k-*medoids, and AHC by testing each of them upon a random selection of participants (1,417). The *k-*modes method used with an alternative initialization technique, called the *Cao* metric ^6^, resulted in some clusters containing less than ten participants, while others contained hundreds. Such imbalance is uninformative for our purposes. Similarly, *k*-medoids with Hamming distance and *Jaccard similarity* ^16^, another similarity metric, resulted in several empty clusters. Finally, AHC with other metrics and linkage types resulted in poor separation between clusters, with high overlap between branches. These issues were not present with the specified metrics.

**Supplementary Appendix 2.** Bubble Heatmap.

The *bubble heatmap* places ARF values on a grid in which the *y-*axis contains conditions, the *x-*axis contains clusters, and data points are colored blue (under-representation) or red (over-representation) at each intersection. The magnitude of under-or over-representation is indicated by the size of the data point, or *bubble.* Points are not statistically significant, as determined by the Fisher’s Exact test (REF), were omitted. Therefore, conditions with no significant values are omitted entirely from the *y-* axis.

Notably, for visualisation purposes, the ARF values are adjusted so that values denoting under-representation (between zero and one) were mapped to a similar scale as those denoting over-representation (values greater than one). Specifically, we used the following function, in which *x* denotes the original ARF value:

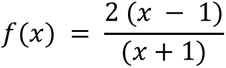

**Supplementary Table 2. ARF values and adjusted p-values per condition and cluster.**

Statistically significant values (p < 0.05) are denoted in bold.

See corresponding excel file (S2Table.xlsx).

**Supplementary Table 2.a.**
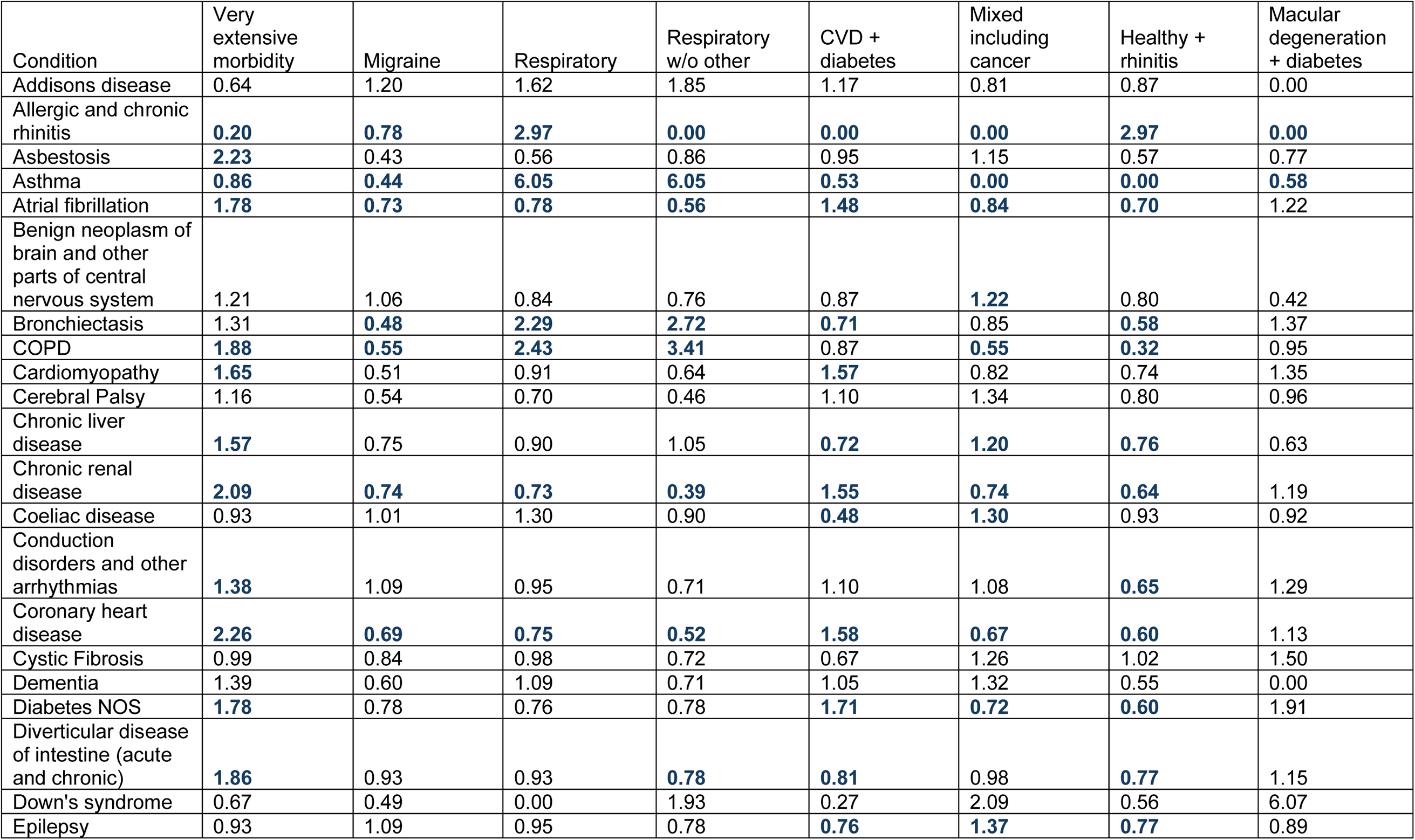
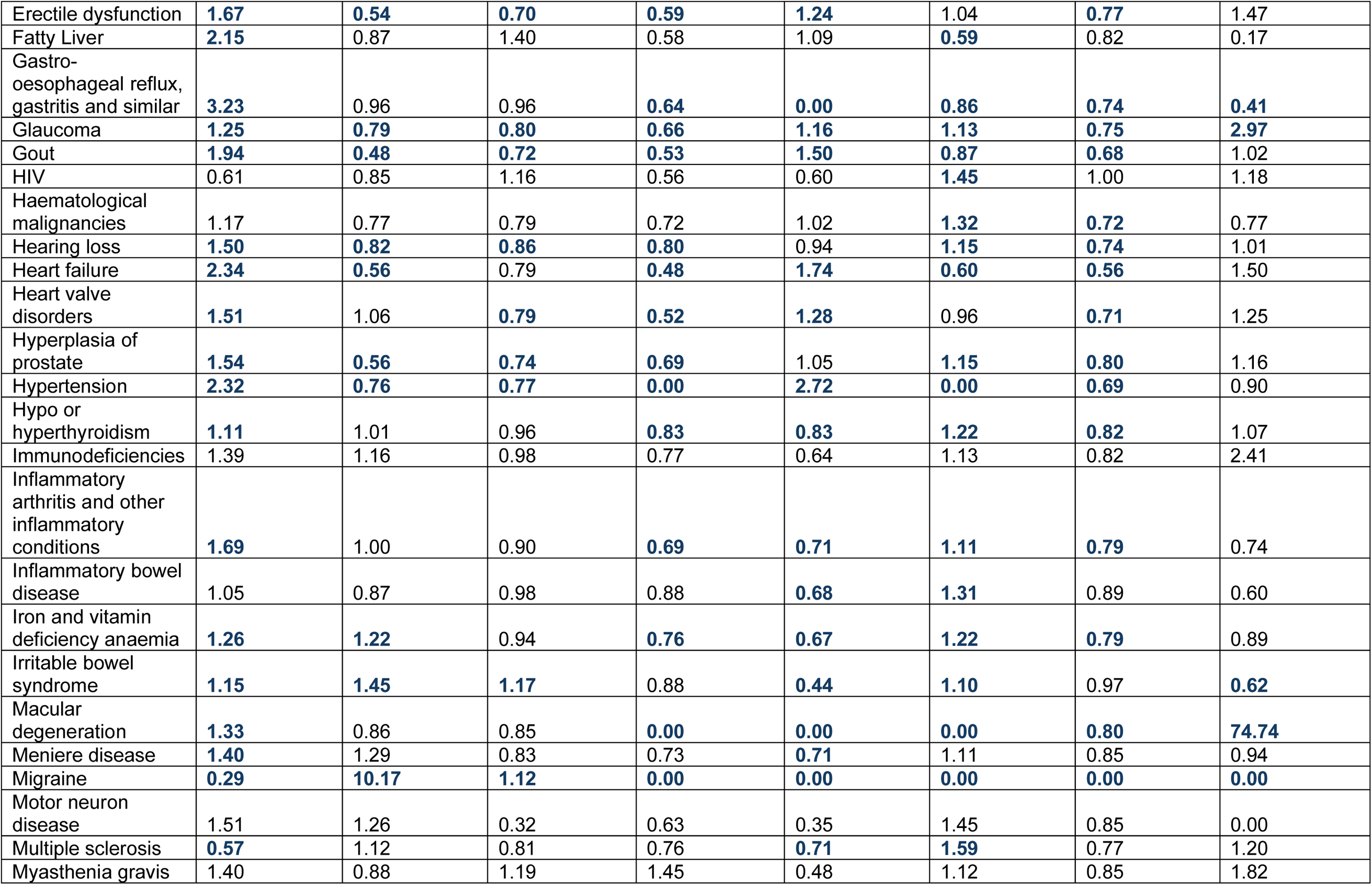
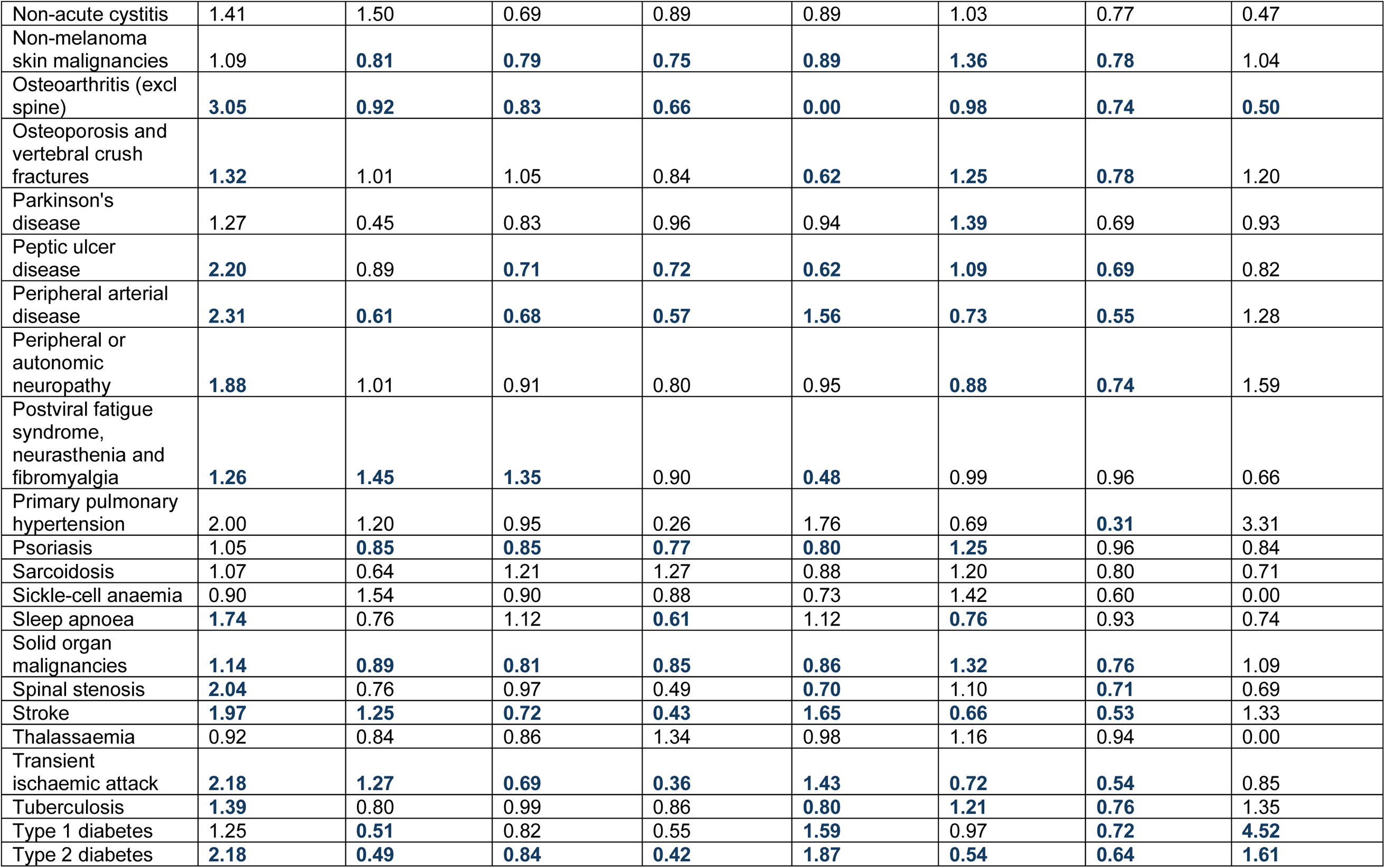
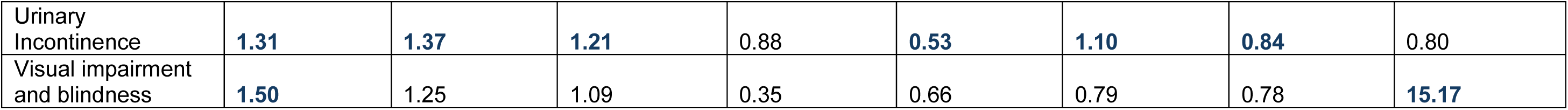
ARF values for *whole*-cohort clusters.

**Supplementary Table 2.b.** ARF values for *women-only* clusters.

| Condition | Very extensive morbidity | Extensive morbidity | Respiratory | Digestive | MSK | CVD + diabetes | Mixed including cancer | Healthy + rhinitis |
| --- | --- | --- | --- | --- | --- | --- | --- | --- |
| Addisons disease | 1.28 | 1.20 | 1.15 | 0.00 | 0.56 | 0.74 | 1.43 | 0.80 |
| Allergic and chronic rhinitis | **2.37** | **0.44** | **2.80** | **0.68** | **0.44** | **0.53** | **0.00** | **2.80** |
| Asbestosis | 0.00 | 0.00 | 0.00 | 0.00 | 0.87 | 2.18 | 0.98 | 0.90 |
| Asthma | **1.67** | **0.74** | **5.79** | **0.58** | **0.54** | **0.47** | **0.78** | **0.00** |
| Atrial fibrillation | **1.91** | 1.42 | **0.63** | 1.54 | **1.25** | **1.50** | **0.76** | **0.29** |
| Benign neoplasm of brain and other parts of central nervous system | 1.27 | 0.84 | 0.83 | 0.94 | 1.07 | 1.02 | 1.17 | **0.63** |
| Bronchiectasis | 1.61 | 1.20 | **2.29** | 0.56 | 1.01 | 0.74 | 0.96 | **0.49** |
| COPD | **2.03** | **1.81** | **2.26** | 2.08 | 1.01 | 0.86 | **0.83** | **0.20** |
| Cardiomyopathy | 2.06 | 1.37 | 1.09 | 1.08 | 0.62 | **1.79** | 0.75 | **0.34** |
| Cerebral Palsy | 0.94 | 1.96 | 0.00 | 2.47 | 1.09 | 0.91 | 1.40 | 0.45 |
| Chronic liver disease | 1.37 | 1.58 | 0.91 | 1.05 | 0.92 | 0.76 | **1.24** | 0.68 |
| Chronic renal disease | **1.67** | **1.77** | **0.65** | 1.96 | 1.08 | **1.63** | **0.70** | **0.36** |
| Coeliac disease | 1.19 | 1.32 | 1.17 | 2.56 | **0.70** | **0.60** | **1.25** | 1.04 |
| Conduction disorders and other arrhythmias | 1.40 | 1.44 | 0.79 | 1.58 | 0.89 | 1.17 | 1.08 | **0.56** |
| Coronary heart disease | **2.21** | **2.04** | **0.68** | **1.88** | 1.10 | **1.66** | **0.57** | **0.25** |
| Cystic Fibrosis | 0.41 | 1.03 | 0.90 | 1.63 | 0.79 | 0.72 | 1.29 | 1.26 |
| Dementia | 1.29 | 1.07 | 0.62 | 0.00 | 1.19 | 0.99 | 1.33 | 0.31 |
| Diabetes NOS | 1.83 | 1.50 | 0.62 | 2.25 | 1.00 | **1.57** | 0.78 | **0.39** |
| Diverticular disease of intestine (acute and chronic) | **2.24** | **2.14** | 0.84 | **1.73** | 1.10 | 0.94 | **0.86** | **0.51** |
| Down's syndrome | 0.00 | 0.00 | 0.00 | 0.00 | 0.32 | 0.27 | **2.69** | 0.66 |
| Epilepsy | 1.16 | 1.04 | 0.98 | 0.48 | 0.92 | **0.77** | **1.33** | **0.73** |
| Erectile dysfunction |  |  |  |  |  |  |  |  |
| Fatty Liver | **3.31** | **3.14** | 0.98 | 1.84 | 0.76 | 1.31 | **0.48** | **0.35** |
| Gastro-oesophageal reflux, gastritis and similar | **1.87** | **5.43** | 1.00 | **5.43** | **0.00** | **0.81** | 0.93 | **0.61** |
| Glaucoma | 1.24 | 1.39 | **0.67** | 0.52 | 1.07 | **1.19** | 1.02 | **0.64** |
| Gout | **1.86** | **1.85** | 0.96 | 2.51 | **1.34** | **1.43** | **0.57** | **0.31** |
| HIV | 0.31 | 0.59 | 1.02 | 0.00 | **0.05** | 0.68 | **1.83** | 1.24 |
| Haematological malignancies | 0.91 | 1.06 | 0.90 | 0.95 | 0.98 | 1.06 | **1.27** | **0.51** |
| Hearing loss | **1.66** | **1.46** | 0.90 | 0.90 | **1.15** | 0.87 | 1.03 | **0.64** |
| Heart failure | **2.19** | 1.56 | 1.03 | 1.76 | 1.20 | **1.65** | **0.56** | **0.16** |
| Heart valve disorders | **1.58** | 1.27 | **0.63** | 1.18 | 0.95 | **1.28** | 1.03 | **0.53** |
| Hyperplasia of prostate |  |  |  |  |  |  |  |  |
| Hypertension | **2.57** | 1.06 | **0.60** | 1.13 | **0.87** | **3.09** | **0.00** | **0.00** |
| Hypo or hyperthyroidism | **1.18** | **1.14** | **0.84** | 0.95 | 0.94 | 1.01 | **1.17** | **0.70** |
| Immunodeficiencies | 1.81 | 0.97 | 0.93 | 4.07 | 0.36 | 1.12 | 1.36 | 0.46 |
| Inflammatory arthritis and other inflammatory conditions | **1.79** | **1.93** | **0.77** | 1.23 | **1.37** | **0.79** | 0.91 | **0.59** |
| Inflammatory bowel disease | 1.21 | 1.22 | 0.84 | 0.98 | 0.89 | **0.74** | **1.29** | 0.84 |
| Iron and vitamin deficiency anaemia | 1.14 | **1.30** | 0.92 | 1.47 | **0.79** | **0.84** | **1.28** | **0.79** |
| Irritable bowel syndrome | **4.46** | 0.85 | **0.69** | **1.39** | **0.47** | **0.49** | **1.11** | **0.90** |
| Macular degeneration | **1.72** | 1.26 | 0.75 | 0.80 | 1.05 | 1.16 | 1.00 | **0.54** |
| Meniere disease | **2.20** | **1.90** | 0.78 | 1.58 | 0.96 | 0.81 | 0.98 | **0.71** |
| Migraine | **1.42** | 1.13 | 0.98 | 1.31 | **0.85** | **0.76** | **1.22** | **0.85** |
| Motor neuron disease | 3.20 | 1.99 | 0.00 | 0.00 | 1.11 | 0.46 | 1.24 | 0.57 |
| Multiple sclerosis | 0.95 | 0.53 | 0.73 | 1.29 | **0.67** | 0.74 | **1.61** | 0.82 |
| Myasthenia gravis | 2.20 | 0.91 | 0.79 | 0.00 | 1.15 | 0.53 | 1.14 | 0.92 |
| Non-acute cystitis | 1.70 | 2.02 | 0.66 | 2.78 | 0.88 | 0.98 | 0.97 | 0.73 |
| Non-melanoma skin malignancies | 0.97 | 1.07 | **0.77** | 1.07 | 1.07 | **0.85** | **1.28** | **0.69** |
| Osteoarthritis (excl spine) | **3.19** | **3.71** | **0.55** | **0.00** | **3.71** | **0.00** | **0.00** | **0.00** |
| Osteoporosis and vertebral crush fractures | **1.39** | **1.53** | 0.90 | 1.27 | **1.20** | **0.77** | **1.09** | **0.64** |
| Parkinson's disease | 1.47 | 1.64 | 0.85 | 2.31 | 1.27 | 0.89 | 1.00 | 0.53 |
| Peptic ulcer disease | **1.71** | **3.43** | 0.80 | **35.75** | **0.69** | **0.43** | **0.61** | **0.30** |
| Peripheral arterial disease | **2.05** | **2.07** | 0.62 | 2.26 | 1.01 | **1.37** | 0.79 | **0.37** |
| Peripheral or autonomic neuropathy | **2.00** | **1.91** | 0.93 | 2.03 | 1.22 | 0.87 | 0.89 | **0.51** |
| Postviral fatigue syndrome, neurasthenia and fibromyalgia | **2.35** | **1.84** | 1.13 | 1.30 | 0.90 | **0.61** | 0.96 | 0.85 |
| Primary pulmonary hypertension | 0.00 | 2.53 | 0.73 | 2.66 | 0.82 | 1.96 | 0.98 | **0.00** |
| Psoriasis | 1.16 | 1.18 | 0.86 | 1.34 | 1.01 | **0.77** | **1.21** | **0.83** |
| Sarcoidosis | 0.99 | 1.71 | 1.15 | 1.30 | 0.99 | 1.05 | 1.02 | 0.61 |
| Sickle-cell anaemia | 0.95 | 0.00 | 1.37 | 0.00 | 1.16 | 0.83 | 1.38 | 0.51 |
| Sleep apnoea | **2.98** | 1.52 | 1.27 | 2.31 | 0.83 | 1.23 | **0.62** | **0.54** |
| Solid organ malignancies | 1.02 | 1.16 | **0.76** | 1.17 | 0.95 | 0.92 | **1.27** | **0.71** |
| Spinal stenosis | **2.51** | **2.40** | 0.91 | 1.06 | **1.94** | **0.56** | **0.63** | **0.37** |
| Stroke | **1.94** | **1.64** | 0.71 | 1.86 | 1.07 | **1.63** | **0.70** | **0.28** |
| Thalassaemia | 0.57 | 1.91 | 0.83 | 0.00 | 0.66 | 0.89 | 1.26 | 1.03 |
| Transient ischaemic attack | **2.17** | **2.21** | **0.53** | 1.33 | 1.11 | **1.53** | **0.67** | **0.29** |
| Tuberculosis | **1.52** | 1.46 | 0.82 | 0.88 | 1.22 | 0.87 | 1.01 | **0.71** |
| Type 1 diabetes | 1.15 | 1.01 | 0.83 | 2.13 | 0.71 | **1.57** | 0.98 | 0.61 |
| Type 2 diabetes | **2.33** | **1.58** | **0.79** | 1.24 | 1.09 | **1.90** | **0.47** | **0.22** |
| Urinary Incontinence | **1.81** | **1.57** | 0.98 | 1.11 | 1.03 | **0.76** | **1.05** | **0.73** |
| Visual impairment and blindness | **2.37** | 1.53 | 0.89 | 0.74 | 1.05 | 0.98 | 0.92 | 0.54 |

**Supplementary Table 2.c.** ARF values for *men-only* clusters.

| Condition | Very extensive morbidity | MSK + others | Respiratory + gout + male genitourinary | Digestive | CVD + diabetes | CVD + CKD + gout | Mixed including cancer | Healthy + rhinitis |
| --- | --- | --- | --- | --- | --- | --- | --- | --- |
| Addisons disease | 1.29 | 0.48 | 0.00 | 0.00 | 1.13 | 0.64 | 0.73 | 1.93 |
| Allergic and chronic rhinitis | **0.49** | **0.80** | **2.78** | **0.71** | **0.74** | **0.38** | **0.00** | **3.20** |
| Asbestosis | 1.17 | **1.66** | 0.54 | 1.85 | 0.75 | 0.58 | 1.05 | **0.44** |
| Asthma | **0.72** | **0.86** | **6.08** | **0.76** | **0.82** | **0.39** | 1.02 | **1.46** |
| Atrial fibrillation | **1.54** | 1.01 | 1.37 | 1.05 | **1.18** | **1.81** | **0.80** | **0.43** |
| Benign neoplasm of brain and other parts of central nervous system | 1.50 | 1.10 | 1.16 | 0.55 | 0.89 | 0.91 | 1.15 | 0.70 |
| Bronchiectasis | 1.04 | 1.16 | 2.20 | 0.55 | 0.83 | 0.94 | 1.23 | 0.81 |
| COPD | **1.30** | **1.44** | **2.82** | **1.48** | 0.91 | 0.96 | 0.85 | **0.46** |
| Cardiomyopathy | 1.61 | 0.89 | 2.60 | 1.02 | 1.14 | **2.15** | 0.68 | **0.55** |
| Cerebral Palsy | 0.44 | 1.14 | 2.06 | 0.88 | 1.12 | 1.10 | 1.40 | 0.42 |
| Chronic liver disease | 1.35 | 0.84 | 1.74 | **2.12** | 0.82 | 1.32 | 1.20 | **0.63** |
| Chronic renal disease | **1.95** | 0.90 | 1.23 | 1.03 | **1.22** | **2.57** | **0.64** | **0.26** |
| Coeliac disease | 0.91 | 0.79 | 2.29 | 0.93 | 0.75 | 0.84 | **1.53** | 0.98 |
| Conduction disorders and other arrhythmias | 1.24 | 0.90 | 0.91 | 1.14 | 1.14 | 1.36 | 1.13 | **0.50** |
| Coronary heart disease | **1.70** | **1.10** | 1.08 | 1.07 | **1.33** | **1.49** | **0.69** | **0.26** |
| Cystic Fibrosis | 0.00 | 0.44 | 0.00 | 2.35 | 1.03 | 5.28 | 0.40 | 1.45 |
| Dementia | 1.68 | 1.47 | 1.74 | 1.48 | 0.87 | 1.48 | 0.51 | 0.61 |
| Diabetes NOS | **2.61** | 0.84 | 1.30 | 0.74 | **1.38** | 1.06 | **0.66** | **0.20** |
| Diverticular disease of intestine (acute and chronic) | **1.26** | **1.29** | 1.64 | 1.30 | 0.89 | **1.29** | 0.99 | **0.52** |
| Down's syndrome | 0.00 | 1.05 | 0.00 | 0.00 | 0.00 | 0.00 | 2.89 | 1.16 |
| Epilepsy | 0.99 | 1.02 | 0.69 | 0.79 | 0.82 | 0.78 | **1.55** | **0.69** |
| Erectile dysfunction | **8.11** | **0.57** | **4.30** | **0.44** | **0.00** | **0.00** | **0.00** | **0.41** |
| Fatty Liver | **1.64** | 1.03 | 1.06 | 1.09 | 1.19 | 1.36 | **0.69** | **0.61** |
| Gastro-oesophageal reflux, gastritis and similar | **1.14** | **1.15** | 1.10 | **2.31** | **0.77** | 0.94 | **1.14** | **0.65** |
| Glaucoma | 1.24 | 0.97 | 1.05 | 0.85 | 1.08 | 1.03 | **1.20** | **0.57** |
| Gout | **1.36** | **0.48** | **8.30** | **0.32** | **0.00** | **13.49** | **0.00** | **0.27** |
| HIV | 1.11 | 0.66 | 1.47 | 1.04 | 0.77 | 0.31 | **1.78** | 0.86 |
| Haematological malignancies | 1.12 | 0.93 | 0.67 | 1.36 | 0.89 | 1.48 | **1.28** | **0.64** |
| Hearing loss | **1.31** | **1.29** | 1.37 | 1.06 | **0.85** | 0.90 | **1.17** | **0.51** |
| Heart failure | **1.82** | 1.03 | 1.40 | 1.22 | **1.29** | **2.29** | **0.55** | **0.24** |
| Heart valve disorders | **1.38** | 1.03 | 1.39 | 1.25 | **1.16** | 1.37 | 0.90 | **0.48** |
| Hyperplasia of prostate | **1.46** | **1.24** | **1.42** | 0.86 | **0.85** | 0.84 | **1.12** | **0.60** |
| Hypertension | **1.57** | **0.86** | **0.60** | **0.80** | **2.37** | **1.70** | **0.00** | **0.00** |
| Hypo or hyperthyroidism | **1.24** | 0.94 | 1.55 | 0.82 | 1.01 | 0.82 | **1.24** | **0.68** |
| Immunodeficiencies | 1.61 | 1.38 | 0.00 | 0.00 | 0.78 | 0.80 | 1.18 | 0.66 |
| Inflammatory arthritis and other inflammatory conditions | 1.02 | **1.62** | 1.33 | 1.04 | **0.75** | 1.31 | 1.02 | **0.51** |
| Inflammatory bowel disease | 1.03 | 0.93 | 0.94 | 1.28 | **0.76** | 0.76 | **1.37** | 0.96 |
| Iron and vitamin deficiency anaemia | **1.30** | 1.09 | 1.40 | **1.83** | **0.79** | 1.08 | 1.14 | **0.63** |
| Irritable bowel syndrome | 0.96 | 1.01 | 1.10 | **1.43** | **0.74** | **0.68** | **1.33** | 0.98 |
| Macular degeneration | **1.44** | 1.01 | 1.38 | 1.02 | 0.99 | 0.91 | 1.17 | **0.56** |
| Meniere disease | 1.02 | 1.22 | 1.54 | 1.39 | 0.87 | 0.82 | 1.31 | **0.48** |
| Migraine | 0.95 | 0.99 | 0.89 | 0.96 | **0.84** | **0.68** | **1.40** | 0.90 |
| Motor neuron disease | 1.23 | 1.81 | 0.00 | 0.00 | 0.48 | 0.81 | 0.93 | 1.12 |
| Multiple sclerosis | **2.32** | **0.43** | 1.03 | 1.17 | 0.73 | 0.44 | **1.52** | 0.78 |
| Myasthenia gravis | 1.67 | 1.31 | 0.00 | 0.88 | 0.90 | 0.00 | 1.05 | 0.73 |
| Non-acute cystitis | 1.52 | 1.08 | 1.57 | 0.45 | 0.98 | 0.67 | 1.38 | 0.37 |
| Non-melanoma skin malignancies | 1.02 | 1.08 | 0.98 | 0.76 | 0.88 | 1.03 | **1.36** | **0.67** |
| Osteoarthritis (excl spine) | 0.95 | **4.45** | 1.19 | **0.00** | **0.00** | 1.01 | **0.00** | **0.00** |
| Osteoporosis and vertebral crush fractures | 1.34 | **1.29** | 1.38 | 0.98 | **0.76** | 0.76 | 1.18 | **0.65** |
| Parkinson's disease | **2.08** | 0.88 | 1.52 | 1.01 | 0.70 | 0.76 | 1.43 | **0.48** |
| Peptic ulcer disease | **0.82** | **1.18** | 0.74 | **18.36** | **0.00** | 0.80 | **0.00** | **0.00** |
| Peripheral arterial disease | **2.29** | 1.14 | 0.47 | 1.32 | **1.20** | 1.33 | **0.61** | **0.20** |
| Peripheral or autonomic neuropathy | **2.02** | **1.30** | 0.99 | 1.36 | 0.87 | 1.16 | **0.78** | **0.44** |
| Postviral fatigue syndrome, neurasthenia and fibromyalgia | 1.20 | 1.05 | 0.99 | 1.12 | **0.64** | 0.77 | **1.26** | 1.07 |
| Primary pulmonary hypertension | 1.48 | 0.87 | 0.00 | 0.78 | 1.84 | 1.17 | 0.67 | 0.16 |
| Psoriasis | 1.08 | 0.94 | 1.03 | 0.84 | **0.82** | 1.01 | **1.34** | 0.89 |
| Sarcoidosis | 0.82 | 0.80 | 2.32 | 0.56 | 0.89 | 0.99 | **1.66** | 0.70 |
| Sickle-cell anaemia | 0.00 | 1.31 | 0.00 | 1.76 | 1.03 | 0.00 | 1.20 | 1.09 |
| Sleep apnoea | **1.62** | **1.21** | 0.95 | 0.95 | 1.02 | 1.34 | **0.80** | **0.57** |
| Solid organ malignancies | **1.62** | 1.00 | 1.27 | 1.07 | **0.83** | 0.99 | **1.25** | **0.57** |
| Spinal stenosis | 1.11 | **2.01** | 1.53 | 0.62 | **0.68** | 1.21 | 0.84 | **0.46** |
| Stroke | **1.73** | 1.02 | 0.61 | 1.07 | **1.44** | **1.67** | **0.64** | **0.23** |
| Thalassaemia | 0.91 | 0.78 | 0.00 | 1.80 | 0.70 | 1.80 | 1.33 | 0.99 |
| Transient ischaemic attack | **1.78** | 1.13 | 0.80 | 0.92 | **1.30** | **1.72** | **0.70** | **0.20** |
| Tuberculosis | 0.98 | 1.01 | 1.89 | 1.07 | 0.84 | 1.13 | **1.47** | **0.56** |
| Type 1 diabetes | **2.92** | **0.53** | 1.72 | 0.59 | 1.23 | 0.22 | 0.99 | **0.42** |
| Type 2 diabetes | **2.56** | 0.91 | 1.10 | 0.84 | **1.37** | **1.57** | **0.49** | **0.22** |
| Urinary Incontinence | **2.20** | 1.24 | 1.55 | 0.79 | **0.63** | 0.48 | 1.03 | 0.71 |
| Visual impairment and blindness | 1.38 | 1.21 | 0.48 | 0.68 | 0.82 | 0.81 | 1.18 | 0.75 |

**Supplementary Table 2.d.** Adjusted *p*-values values for *whole*-cohort clusters.

| Condition | Very extensive morbidity | Migraine | Respiratory | Respiratory w/o other | CVD + diabetes | Mixed including cancer | Healthy + rhinitis | Macular degeneration + diabetes |
| --- | --- | --- | --- | --- | --- | --- | --- | --- |
| Addisons disease | 1.00E+00 | 1.00E+00 | 1.00E+00 | 1.00E+00 | 1.00E+00 | 1.00E+00 | 1.00E+00 | 1.00E+00 |
| Allergic and chronic rhinitis | 0.00E+00 | 5.20E-58 | 0.00E+00 | 0.00E+00 | 0.00E+00 | 0.00E+00 | 0.00E+00 | 3.40E-173 |
| Asbestosis | 1.00E-06 | 8.97E-01 | 1.00E+00 | 1.00E+00 | 1.00E+00 | 1.00E+00 | 7.88E-02 | 1.00E+00 |
| Asthma | 1.90E-14 | 2.34E-213 | 0.00E+00 | 0.00E+00 | 1.40E-270 | 0.00E+00 | 0.00E+00 | 5.89E-08 |
| Atrial fibrillation | 4.36E-53 | 5.07E-05 | 6.81E-03 | 4.06E-07 | 3.04E-28 | 6.59E-07 | 1.16E-19 | 1.00E+00 |
| Benign neoplasm of brain and other parts of central nervous system | 1.00E+00 | 1.00E+00 | 1.00E+00 | 1.00E+00 | 1.00E+00 | 2.99E-02 | 3.17E-01 | 1.00E+00 |
| Bronchiectasis | 5.93E-02 | 3.71E-05 | 1.07E-21 | 2.19E-17 | 1.42E-02 | 4.47E-01 | 2.46E-10 | 1.00E+00 |
| COPD | 3.07E-67 | 2.35E-15 | 9.73E-102 | 8.51E-120 | 1.29E-01 | 6.00E-62 | 1.24E-118 | 1.00E+00 |
| Cardiomyopathy | 1.76E-04 | 7.04E-02 | 1.00E+00 | 1.00E+00 | 1.29E-04 | 1.00E+00 | 2.53E-01 | 1.00E+00 |
| Cerebral Palsy | 1.00E+00 | 1.00E+00 | 1.00E+00 | 1.00E+00 | 1.00E+00 | 7.57E-01 | 1.00E+00 | 1.00E+00 |
| Chronic liver disease | 2.75E-10 | 6.49E-01 | 1.00E+00 | 1.00E+00 | 1.67E-03 | 2.50E-03 | 9.20E-04 | 1.00E+00 |
| Chronic renal disease | 2.86E-113 | 4.51E-05 | 1.96E-05 | 4.12E-17 | 2.02E-41 | 1.88E-21 | 7.15E-33 | 1.00E+00 |
| Coeliac disease | 1.00E+00 | 1.00E+00 | 4.77E-01 | 1.00E+00 | 5.55E-12 | 1.18E-06 | 1.00E+00 | 1.00E+00 |
| Conduction disorders and other arrhythmias | 7.24E-05 | 1.00E+00 | 1.00E+00 | 1.00E+00 | 1.00E+00 | 1.00E+00 | 3.14E-10 | 1.00E+00 |
| Coronary heart disease | 0.00E+00 | 4.70E-28 | 5.24E-17 | 9.89E-36 | 1.39E-171 | 7.11E-119 | 5.50E-137 | 1.00E+00 |
| Cystic Fibrosis | 1.00E+00 | 1.00E+00 | 1.00E+00 | 1.00E+00 | 1.00E+00 | 1.00E+00 | 1.00E+00 | 1.00E+00 |
| Dementia | 1.00E+00 | 1.00E+00 | 1.00E+00 | 1.00E+00 | 1.00E+00 | 1.00E+00 | 1.00E+00 | 1.00E+00 |
| Diabetes NOS | 6.64E-11 | 1.00E+00 | 1.00E+00 | 1.00E+00 | 7.33E-12 | 2.15E-04 | 3.84E-07 | 1.00E+00 |
| Diverticular disease of intestine (acute and chronic) | 9.34E-139 | 1.00E+00 | 1.00E+00 | 5.54E-03 | 2.26E-09 | 1.00E+00 | 1.90E-23 | 1.00E+00 |
| Down's syndrome | 1.00E+00 | 1.00E+00 | 1.00E+00 | 1.00E+00 | 1.00E+00 | 1.44E-01 | 1.00E+00 | 1.00E+00 |
| Epilepsy | 1.00E+00 | 1.00E+00 | 1.00E+00 | 1.00E+00 | 2.71E-05 | 1.06E-26 | 7.46E-08 | 1.00E+00 |
| Erectile dysfunction | 2.55E-104 | 3.35E-41 | 1.45E-15 | 1.77E-15 | 3.70E-19 | 2.46E-01 | 2.32E-27 | 5.01E-02 |
| Fatty Liver | 9.12E-28 | 1.00E+00 | 6.44E-02 | 4.42E-01 | 1.00E+00 | 3.28E-13 | 3.21E-01 | 1.00E+00 |
| Gastro-oesophageal reflux, gastritis and similar | 0.00E+00 | 1.00E+00 | 1.00E+00 | 5.25E-44 | 0.00E+00 | 7.78E-28 | 4.08E-119 | 1.70E-20 |
| Glaucoma | 6.27E-06 | 9.19E-03 | 2.85E-02 | 6.19E-04 | 4.75E-03 | 4.24E-05 | 9.70E-14 | 5.77E-13 |
| Gout | 7.73E-131 | 2.32E-37 | 4.51E-09 | 2.28E-14 | 2.46E-52 | 7.52E-07 | 3.29E-37 | 1.00E+00 |
| HIV | 1.00E+00 | 1.00E+00 | 1.00E+00 | 1.00E+00 | 4.24E-01 | 1.27E-03 | 1.00E+00 | 1.00E+00 |
| Haematological malignancies | 1.00E+00 | 6.07E-01 | 1.00E+00 | 1.00E+00 | 1.00E+00 | 4.93E-11 | 8.21E-07 | 1.00E+00 |
| Hearing loss | 2.12E-46 | 3.62E-04 | 4.36E-02 | 3.38E-02 | 1.00E+00 | 6.38E-14 | 1.69E-27 | 1.00E+00 |
| Heart failure | 1.14E-63 | 2.80E-06 | 1.00E+00 | 3.33E-04 | 2.28E-27 | 1.94E-21 | 2.26E-19 | 1.00E+00 |
| Heart valve disorders | 6.49E-20 | 1.00E+00 | 3.89E-02 | 4.96E-07 | 6.86E-08 | 1.00E+00 | 2.83E-15 | 1.00E+00 |
| Hyperplasia of prostate | 1.91E-56 | 1.02E-30 | 3.24E-09 | 2.32E-06 | 1.00E+00 | 6.54E-14 | 6.21E-16 | 1.00E+00 |
| Hypertension | 0.00E+00 | 1.04E-85 | 5.84E-71 | 0.00E+00 | 0.00E+00 | 0.00E+00 | 0.00E+00 | 1.00E+00 |
| Hypo or hyperthyroidism | 5.80E-06 | 1.00E+00 | 1.00E+00 | 3.97E-03 | 1.52E-14 | 1.96E-65 | 2.95E-22 | 1.00E+00 |
| Immunodeficiencies | 1.00E+00 | 1.00E+00 | 1.00E+00 | 1.00E+00 | 1.00E+00 | 1.00E+00 | 1.00E+00 | 1.00E+00 |
| Inflammatory arthritis and other inflammatory conditions | 7.01E-77 | 1.00E+00 | 1.00E+00 | 1.29E-05 | 1.15E-21 | 1.71E-06 | 9.72E-16 | 1.00E+00 |
| Inflammatory bowel disease | 1.00E+00 | 1.00E+00 | 1.00E+00 | 1.00E+00 | 1.49E-11 | 3.32E-22 | 2.09E-01 | 1.00E+00 |
| Iron and vitamin deficiency anaemia | 4.11E-16 | 6.14E-07 | 1.00E+00 | 2.97E-04 | 1.33E-35 | 8.77E-36 | 3.23E-19 | 1.00E+00 |
| Irritable bowel syndrome | 1.70E-12 | 7.68E-57 | 2.34E-09 | 1.71E-01 | 3.08E-214 | 1.51E-20 | 1.00E+00 | 4.78E-03 |
| Macular degeneration | 8.53E-06 | 1.00E+00 | 1.00E+00 | 3.79E-35 | 1.90E-138 | 1.02E-270 | 1.31E-04 | 0.00E+00 |
| Meniere disease | 1.37E-04 | 3.15E-01 | 1.00E+00 | 1.00E+00 | 1.18E-03 | 1.00E+00 | 7.08E-01 | 1.00E+00 |
| Migraine | 3.69136621242e-312 | 0.00E+00 | 7.75E-05 | 4.04E-280 | 0.00E+00 | 0.00E+00 | 0.00E+00 | 1.19E-42 |
| Motor neuron disease | 1.00E+00 | 1.00E+00 | 1.00E+00 | 1.00E+00 | 1.00E+00 | 1.00E+00 | 1.00E+00 | 1.00E+00 |
| Multiple sclerosis | 2.03E-04 | 1.00E+00 | 1.00E+00 | 1.00E+00 | 3.30E-02 | 9.49E-20 | 5.51E-02 | 1.00E+00 |
| Myasthenia gravis | 1.00E+00 | 1.00E+00 | 1.00E+00 | 1.00E+00 | 1.00E+00 | 1.00E+00 | 1.00E+00 | 1.00E+00 |
| Non-acute cystitis | 6.79E-01 | 7.45E-01 | 1.00E+00 | 1.00E+00 | 1.00E+00 | 1.00E+00 | 1.00E+00 | 1.00E+00 |
| Non-melanoma skin malignancies | 7.32E-01 | 9.62E-04 | 5.58E-05 | 2.55E-03 | 3.77E-02 | 2.71E-62 | 7.53E-17 | 1.00E+00 |
| Osteoarthritis (excl spine) | 0.00E+00 | 8.17E-03 | 1.94E-24 | 1.21E-54 | 0.00E+00 | 2.03E-08 | 1.08E-153 | 5.19E-20 |
| Osteoporosis and vertebral crush fractures | 1.74E-15 | 1.00E+00 | 1.00E+00 | 1.00E+00 | 1.10E-32 | 4.49E-28 | 4.10E-16 | 1.00E+00 |
| Parkinson's disease | 1.00E+00 | 1.11E-01 | 1.00E+00 | 1.00E+00 | 1.00E+00 | 2.10E-03 | 1.88E-01 | 1.00E+00 |
| Peptic ulcer disease | 4.57E-216 | 9.20E-01 | 4.88E-10 | 2.31E-04 | 2.08E-37 | 2.07E-04 | 6.11E-38 | 1.00E+00 |
| Peripheral arterial disease | 1.02E-67 | 3.78E-05 | 4.44E-03 | 6.16E-03 | 7.62E-18 | 4.61E-10 | 8.13E-23 | 1.00E+00 |
| Peripheral or autonomic neuropathy | 3.17E-47 | 1.00E+00 | 1.00E+00 | 1.00E+00 | 1.00E+00 | 1.85E-02 | 4.69E-10 | 1.00E+00 |
| Postviral fatigue syndrome, neurasthenia and fibromyalgia | 2.63E-07 | 3.09E-13 | 5.80E-08 | 1.00E+00 | 4.27E-47 | 1.00E+00 | 1.00E+00 | 1.00E+00 |
| Primary pulmonary hypertension | 1.20E-01 | 1.00E+00 | 1.00E+00 | 1.00E+00 | 3.13E-01 | 1.00E+00 | 1.12E-02 | 1.00E+00 |
| Psoriasis | 1.00E+00 | 1.23E-02 | 2.74E-02 | 3.60E-03 | 2.60E-10 | 1.22E-37 | 1.00E+00 | 1.00E+00 |
| Sarcoidosis | 1.00E+00 | 3.37E-01 | 1.00E+00 | 1.00E+00 | 1.00E+00 | 1.13E-01 | 6.02E-01 | 1.00E+00 |
| Sickle-cell anaemia | 1.00E+00 | 1.00E+00 | 1.00E+00 | 1.00E+00 | 1.00E+00 | 1.00E+00 | 1.00E+00 | 1.00E+00 |
| Sleep apnoea | 2.00E-26 | 1.25E-01 | 1.00E+00 | 1.78E-02 | 1.00E+00 | 4.34E-09 | 1.00E+00 | 1.00E+00 |
| Solid organ malignancies | 1.28E-08 | 2.36E-02 | 2.27E-09 | 1.78E-02 | 1.31E-08 | 6.29E-129 | 9.89E-45 | 1.00E+00 |
| Spinal stenosis | 3.18E-23 | 1.00E+00 | 1.00E+00 | 5.99E-02 | 6.10E-03 | 1.00E+00 | 2.59E-04 | 1.00E+00 |
| Stroke | 5.88E-82 | 8.94E-04 | 2.38E-05 | 4.64E-13 | 3.73E-50 | 1.11E-33 | 6.00E-52 | 1.00E+00 |
| Thalassaemia | 1.00E+00 | 1.00E+00 | 1.00E+00 | 1.00E+00 | 1.00E+00 | 1.00E+00 | 1.00E+00 | 1.00E+00 |
| Transient<br>ischaemic attack | 3.56E-72 | 1.77E-02 | 4.14E-04 | 1.71E-10 | 7.73E-14 | 9.68E-15 | 1.74E-31 | 1.00E+00 |
| Tuberculosis | 2.44E-07 | 5.74E-01 | 1.00E+00 | 1.00E+00 | 2.71E-02 | 5.21E-06 | 2.43E-06 | 1.00E+00 |
| Type 1 diabetes | 1.00E+00 | 1.51E-02 | 1.00E+00 | 1.00E+00 | 1.98E-06 | 1.00E+00 | 2.06E-02 | 2.67E-05 |
| Type 2 diabetes | 1.83E-302 | 6.56E-52 | 6.79E-04 | 7.88E-35 | 2.13E-226 | 7.51E-160 | 8.32E-70 | 8.81E-04 |
| Urinary<br>Incontinence | 5.02E-23 | 5.95E-20 | 2.53E-06 | 1.00E+00 | 8.15E-79 | 8.00E-08 | 9.51E-11 | 1.00E+00 |
| Visual impairment<br>and blindness | 2.88E-02 | 1.00E+00 | 1.00E+00 | 2.66E-01 | 1.82E-01 | 6.64E-01 | 1.00E+00 | 5.24E-33 |

**Supplementary Table 2.e.**
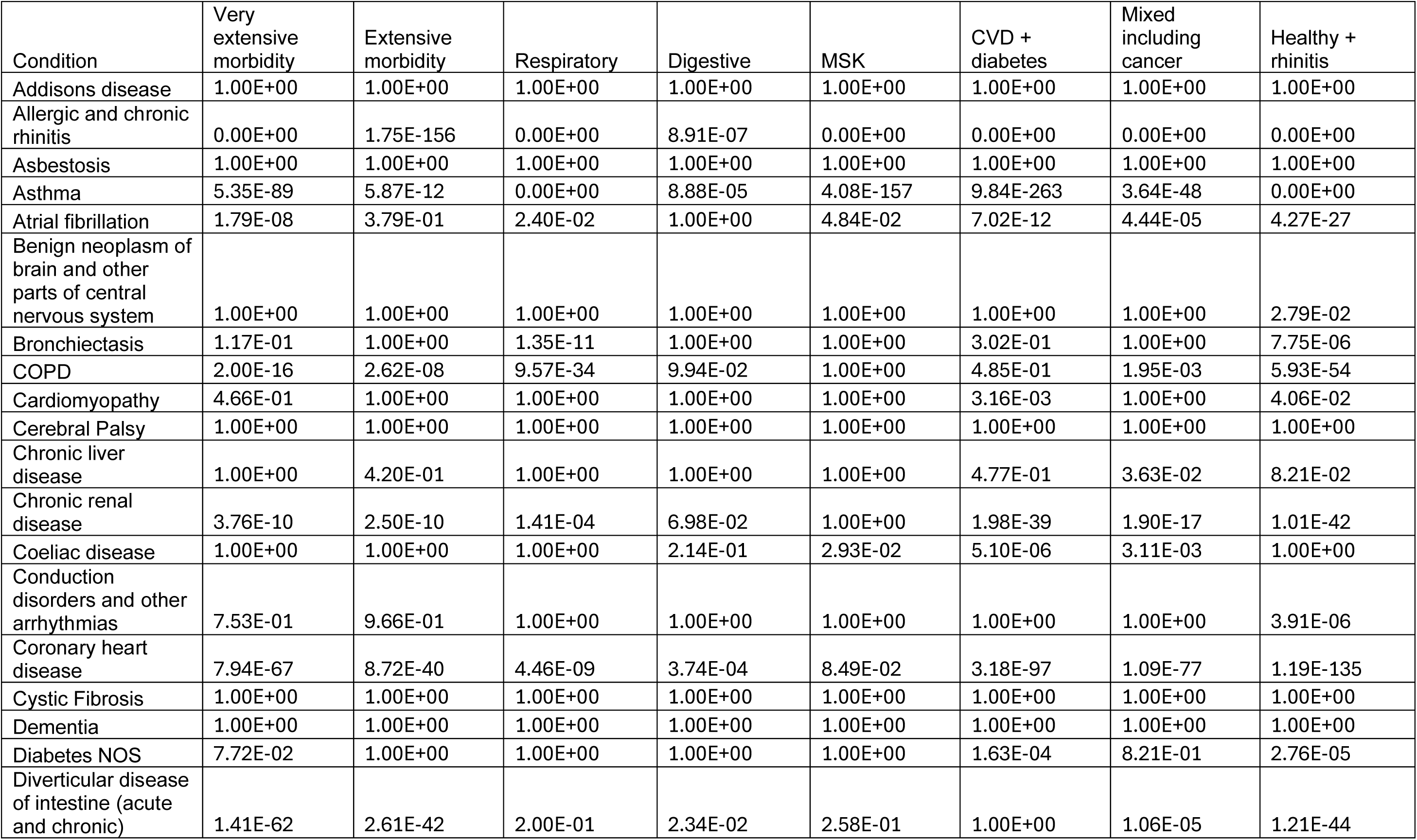
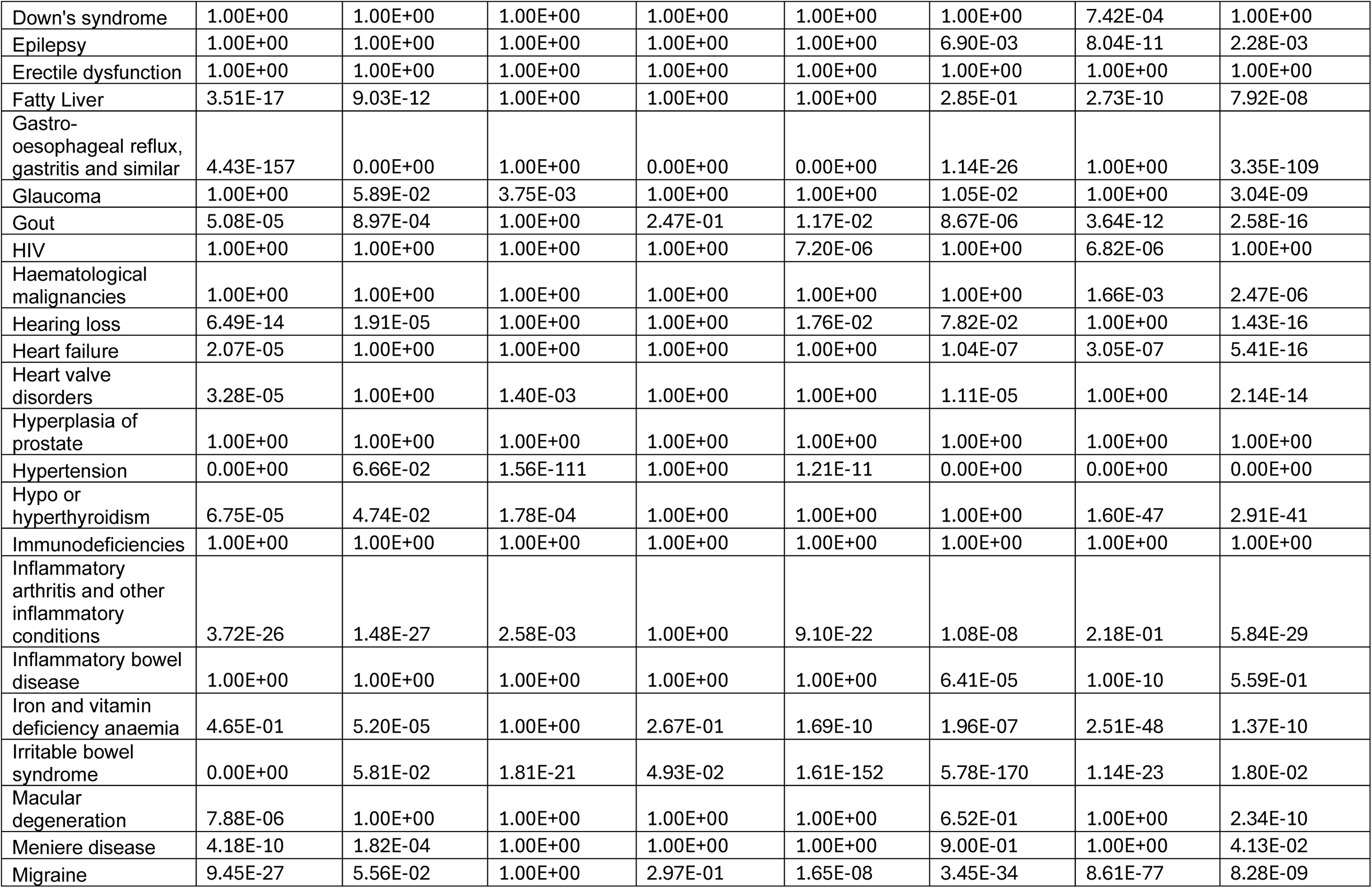
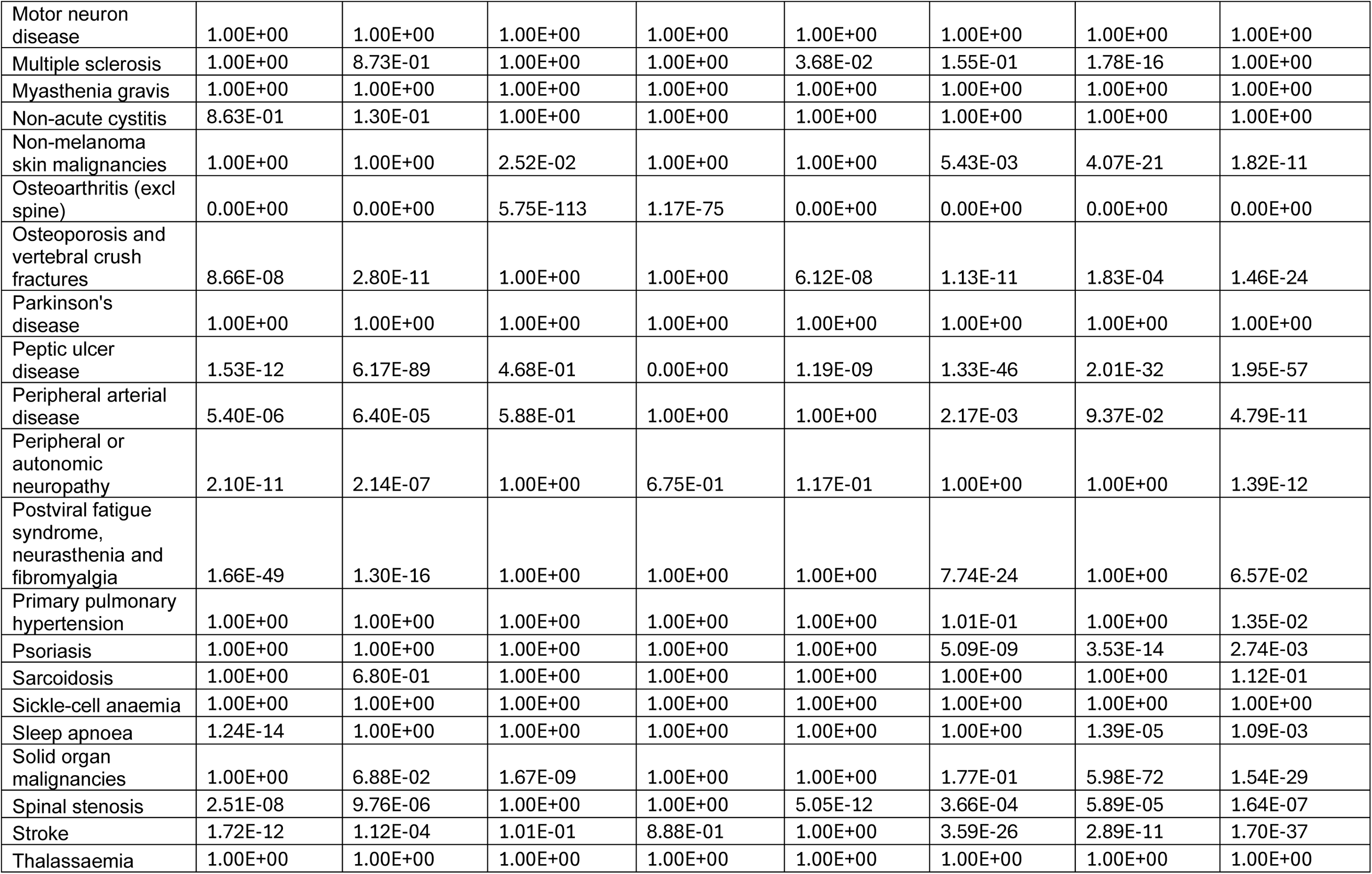
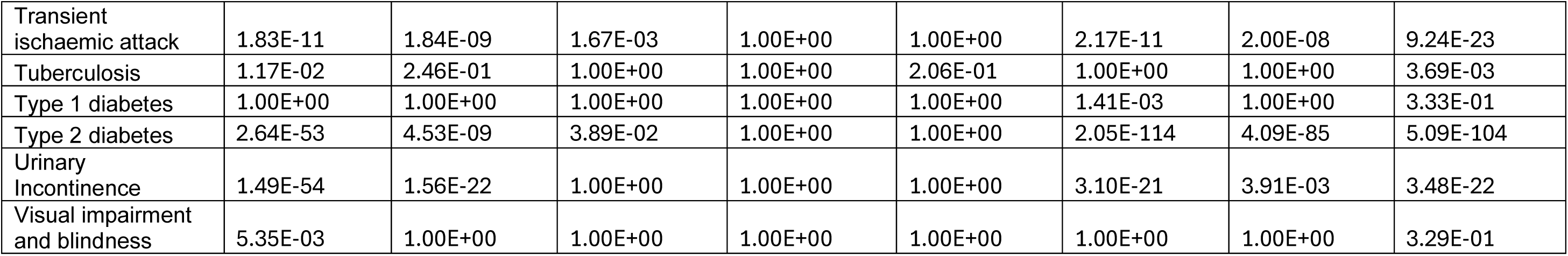
Adjusted *p*-values values for *women-only* clusters.

**Supplementary Table 2.f.**
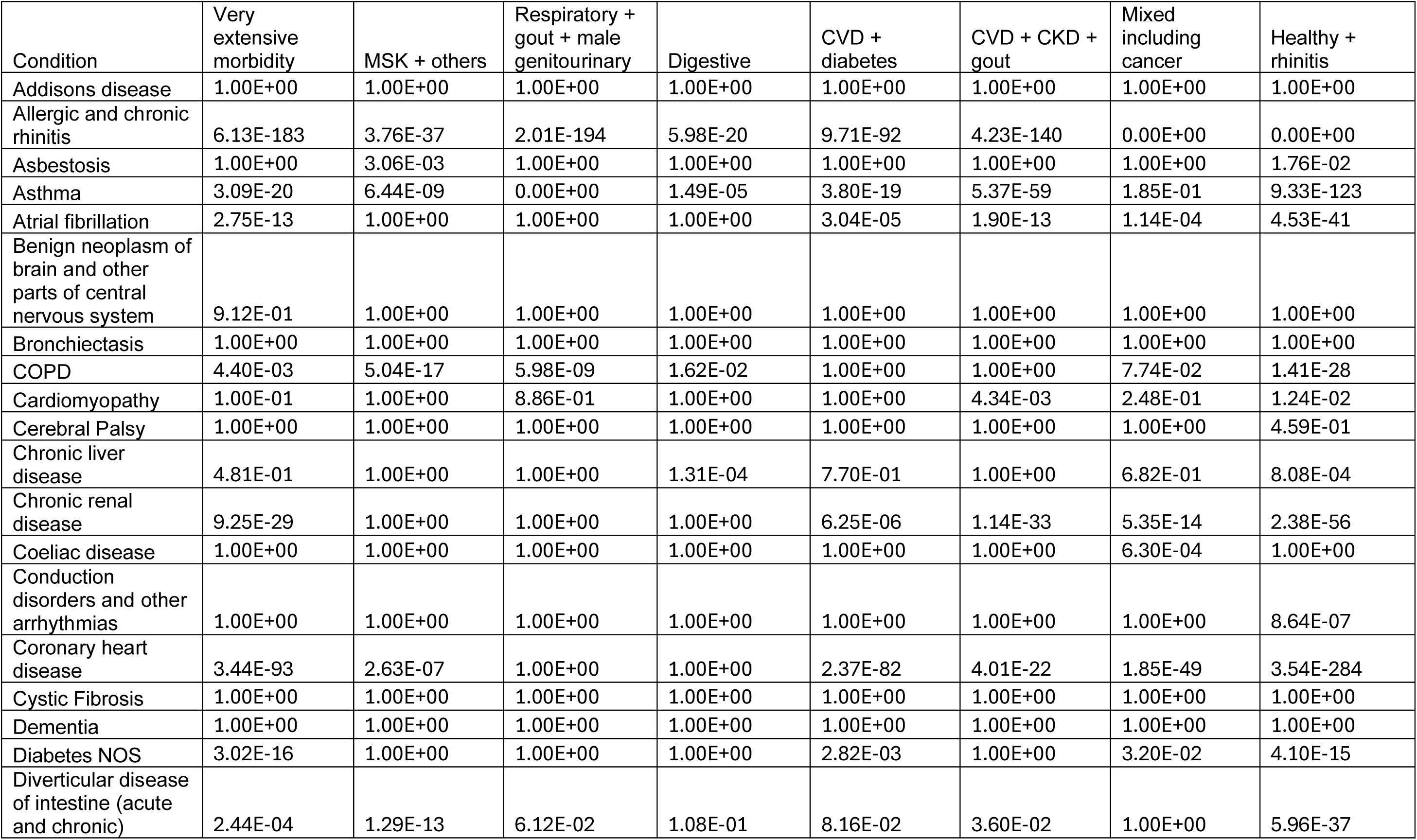
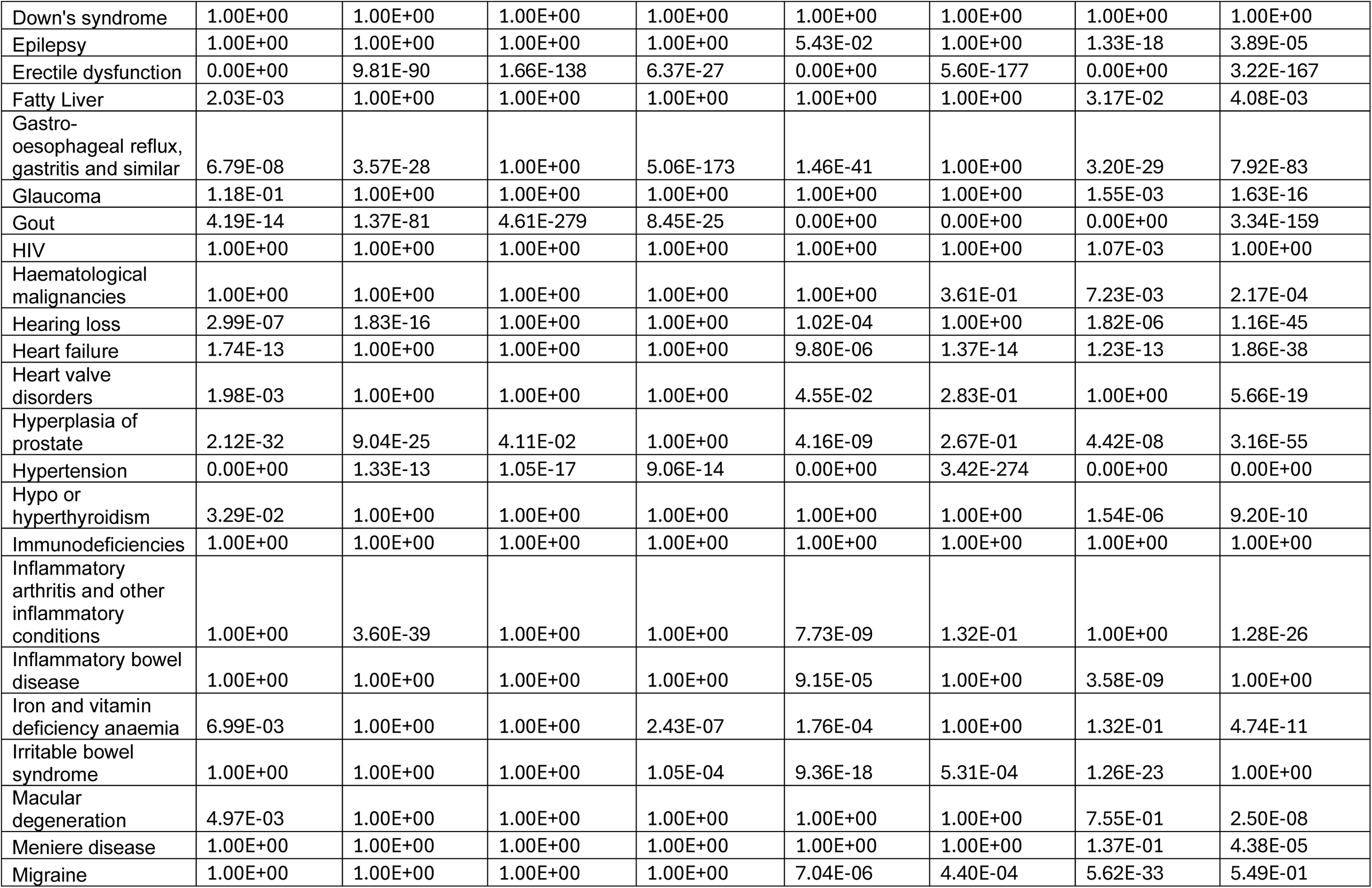
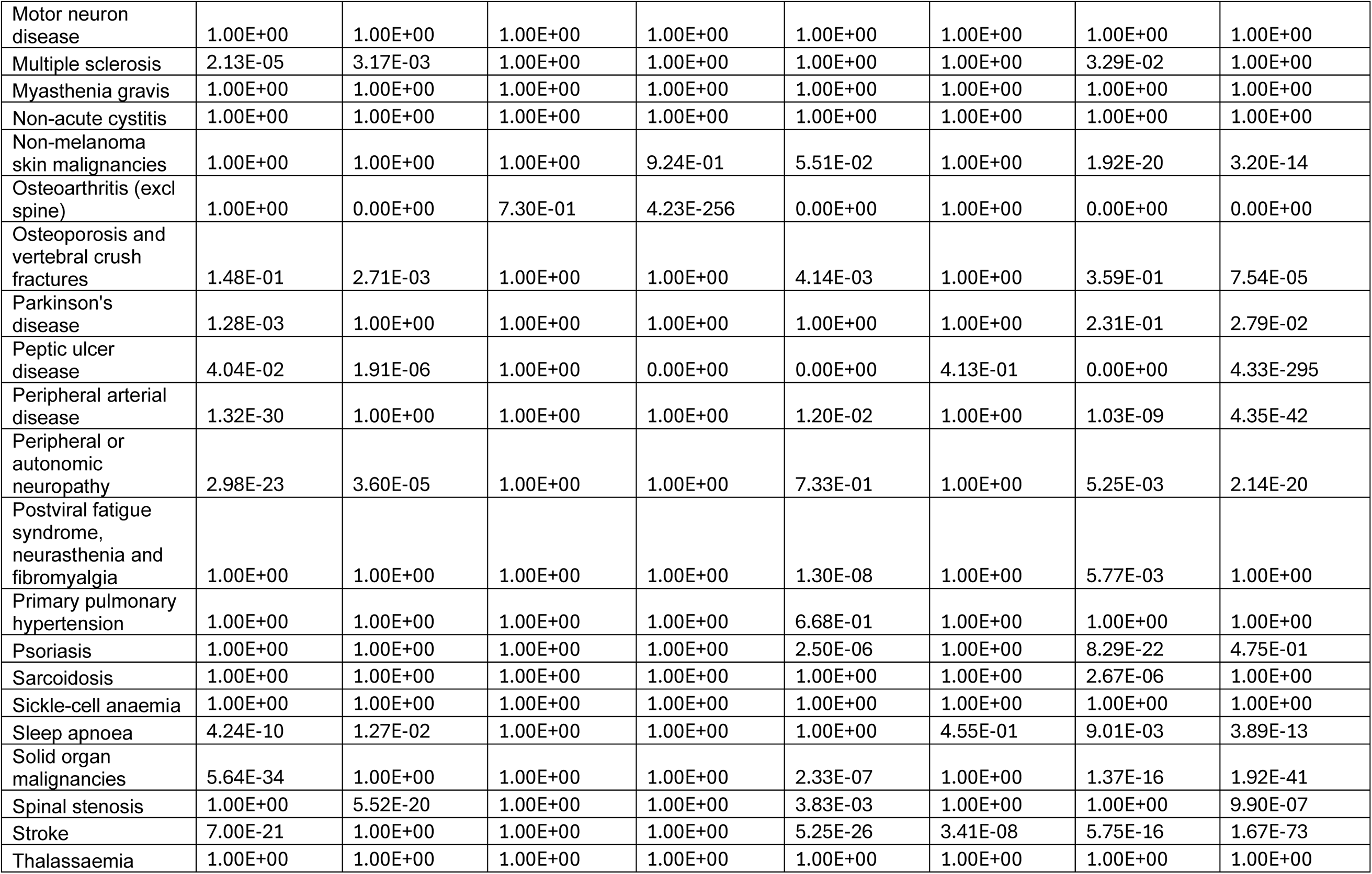
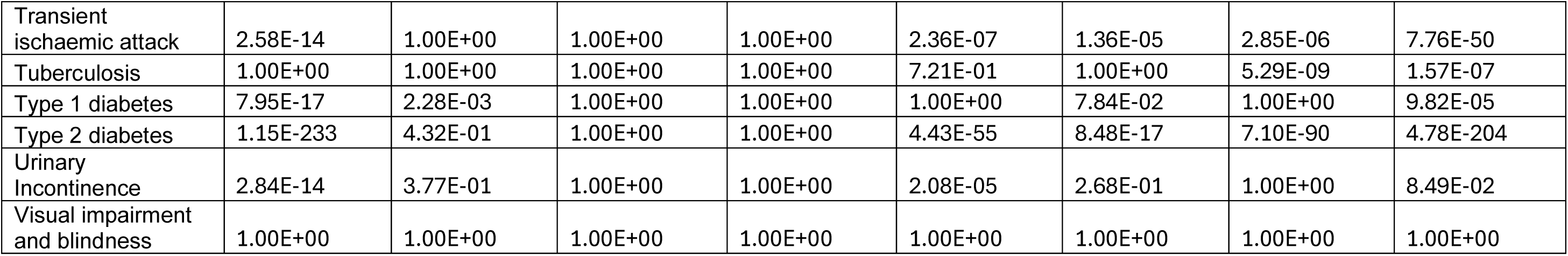
Adjusted *p*-values values for *men-only* clusters.

**Supplementary Table 3.**
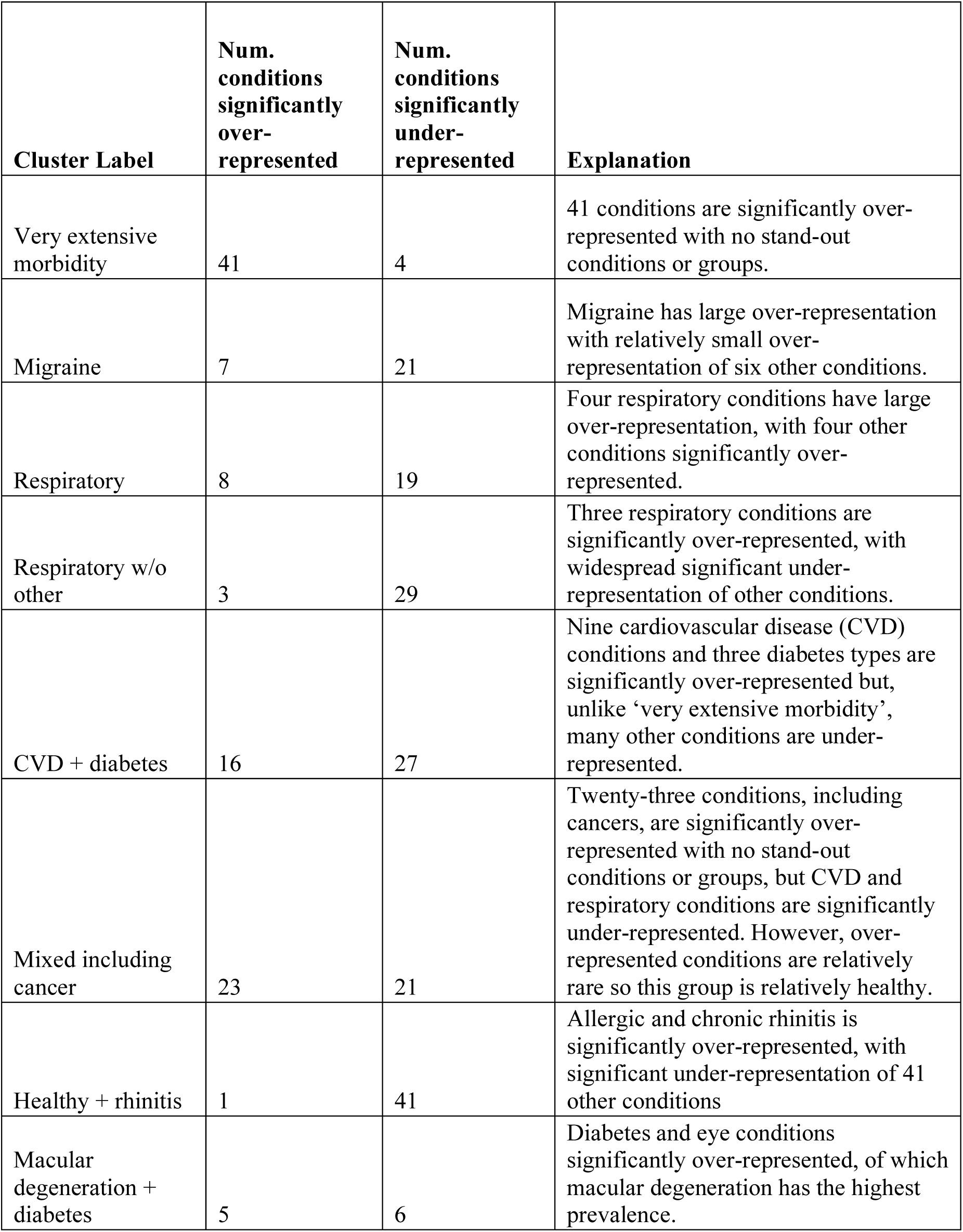
Cluster labels for the *whole* cohort.

| Cluster Label | Num.<br>conditions<br>significantly<br>over-represented | Num.<br>conditions<br>significantly<br>under-represented | Explanation |
| --- | --- | --- | --- |
| Very extensive morbidity | 41 | 4 | 41 conditions are significantly over-represented with no stand-out conditions or groups. |
| Migraine | 7 | 21 | Migraine has large over-representation with relatively small over-representation of six other conditions. |
| Respiratory | 8 | 19 | Four respiratory conditions have large over-representation, with four other conditions significantly over-represented. |
| Respiratory w/o other | 3 | 29 | Three respiratory conditions are significantly over-represented, with widespread significant under-representation of other conditions. |
| CVD + diabetes | 16 | 27 | Nine cardiovascular disease (CVD) conditions and three diabetes types are significantly over-represented but, unlike ‘very extensive morbidity’, many other conditions are under-represented. |
| Mixed including cancer | 23 | 21 | Twenty-three conditions, including cancers, are significantly over-represented with no stand-out conditions or groups, but CVD and respiratory conditions are significantly under-represented. However, over-represented conditions are relatively rare so this group is relatively healthy. |
| Healthy + rhinitis | 1 | 41 | Allergic and chronic rhinitis is significantly over-represented, with significant under-representation of 41 other conditions |
| Macular degeneration + diabetes | 5 | 6 | Diabetes and eye conditions significantly over-represented, of which macular degeneration has the highest prevalence. |

**Supplementary Table 4.**
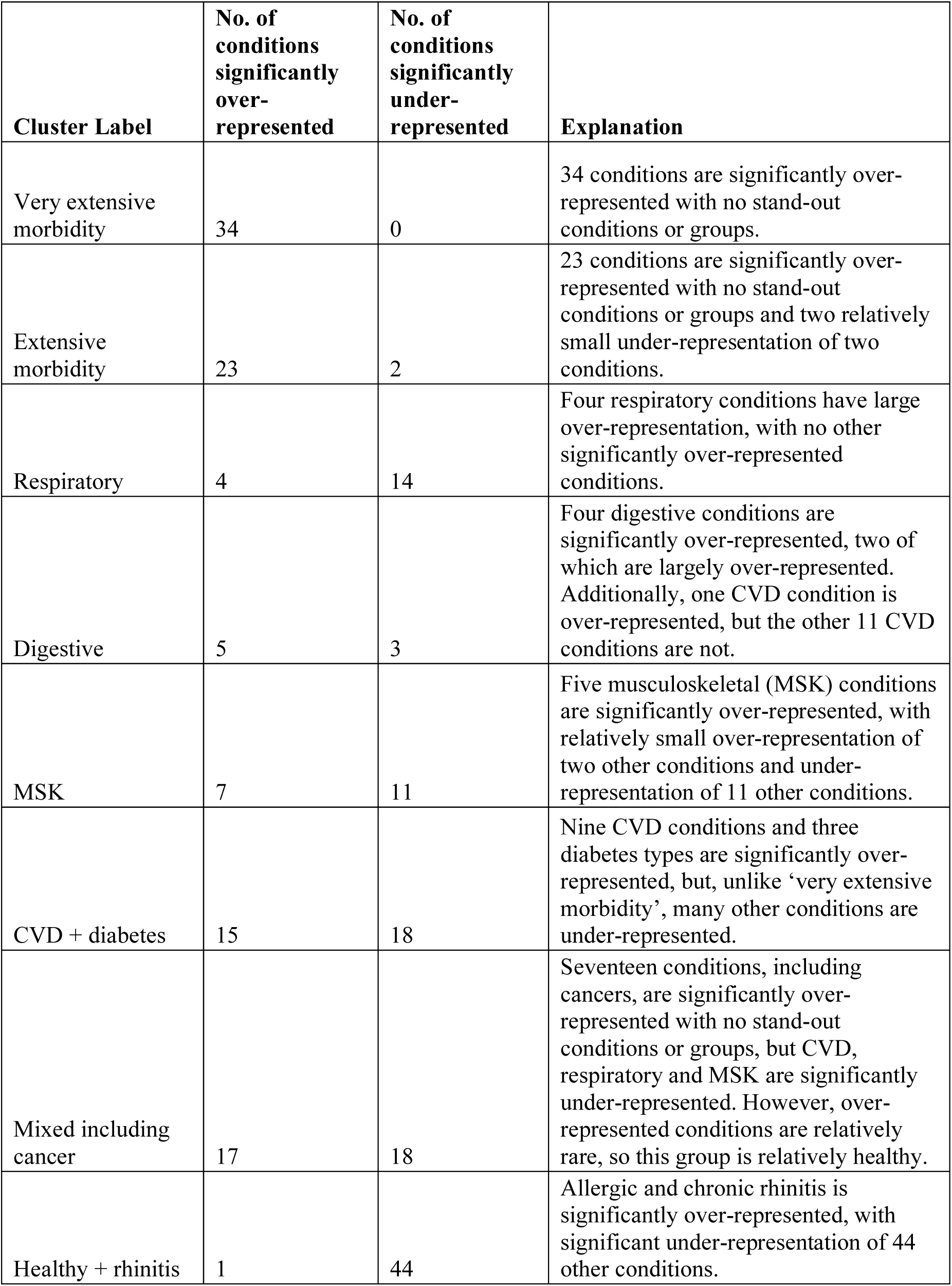
Cluster labels for the *women-only* cohort.

| Cluster Label | No. of conditions significantly over-represented | No. of conditions significantly under-represented | Explanation |
| --- | --- | --- | --- |
| Very extensive morbidity | 34 | 0 | 34 conditions are significantly over-represented with no stand-out conditions or groups. |
| Extensive morbidity | 23 | 2 | 23 conditions are significantly over-represented with no stand-out conditions or groups and two relatively small under-representation of two conditions. |
| Respiratory | 4 | 14 | Four respiratory conditions have large over-representation, with no other significantly over-represented conditions. |
| Digestive | 5 | 3 | Four digestive conditions are significantly over-represented, two of which are largely over-represented. Additionally, one CVD condition is over-represented, but the other 11 CVD conditions are not. |
| MSK | 7 | 11 | Five musculoskeletal (MSK) conditions are significantly over-represented, with relatively small over-representation of two other conditions and under-representation of 11 other conditions. |
| CVD + diabetes | 15 | 18 | Nine CVD conditions and three diabetes types are significantly over-represented, but, unlike 'very extensive morbidity', many other conditions are under-represented. |
| Mixed including cancer | 17 | 18 | Seventeen conditions, including cancers, are significantly over-represented with no stand-out conditions or groups, but CVD, respiratory and MSK are significantly under-represented. However, over-represented conditions are relatively rare, so this group is relatively healthy. |
| Healthy + rhinitis | 1 | 44 | Allergic and chronic rhinitis is significantly over-represented, with significant under-representation of 44 other conditions. |

**Supplementary Table 5.**
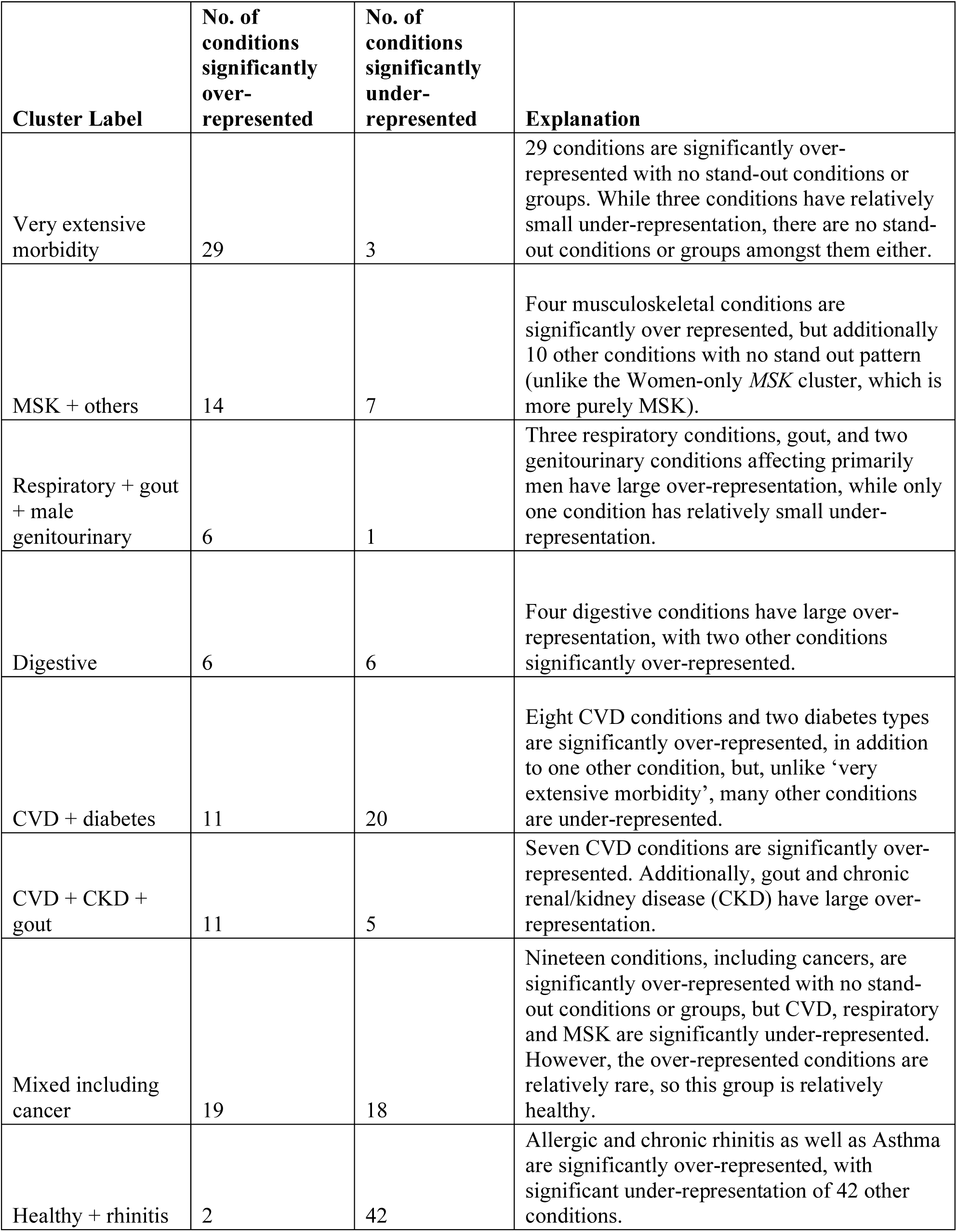
Cluster labels for the *men-only* cohort.

**Supplementary Figure 2.**
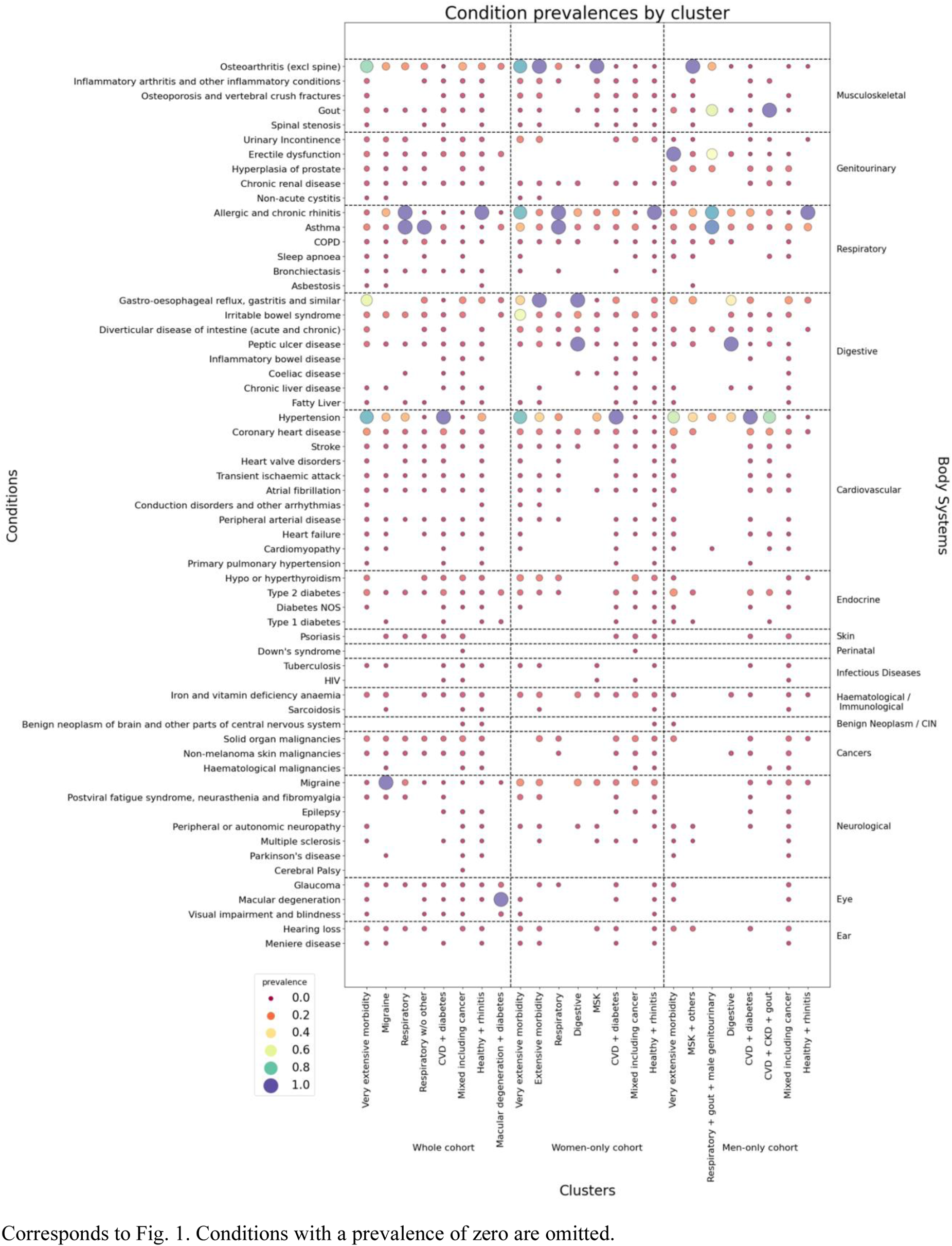
Prevalence values per cluster and condition.

**Supplementary Figure 3.**
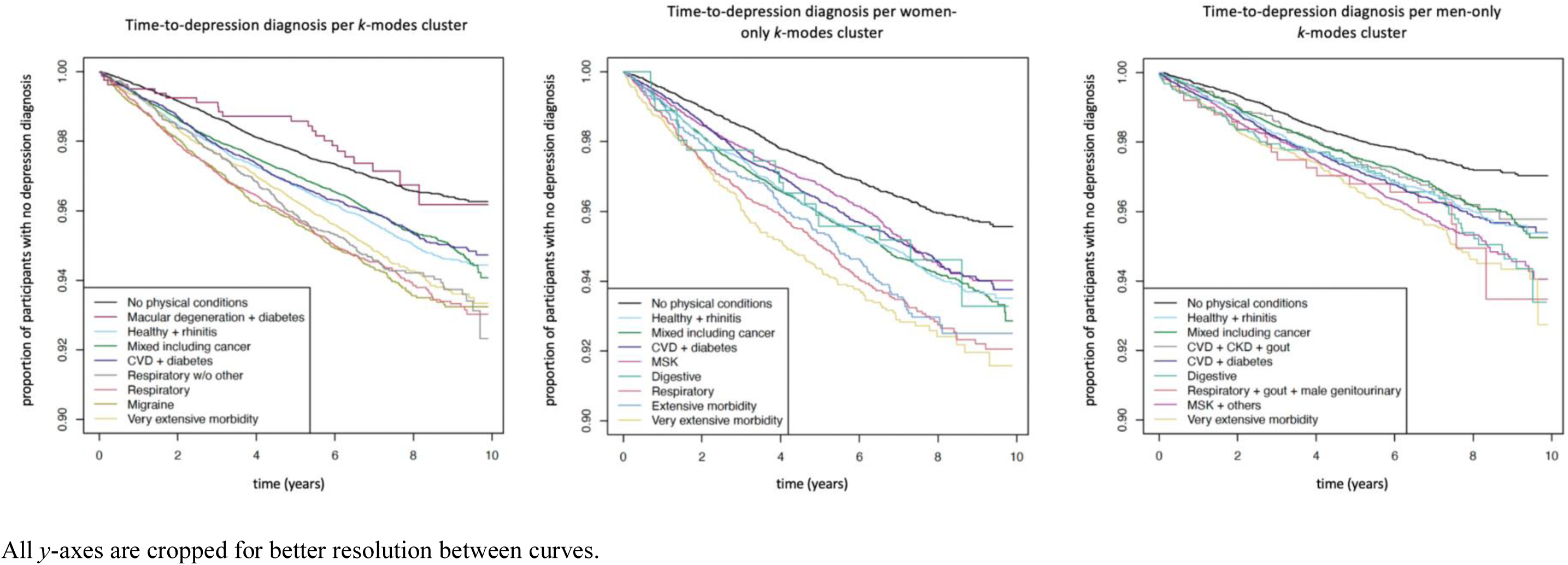
Time-to-depression diagnosis for each cluster in each of the *k*-modes models.

