## Supplementary figures and images for "Investigating associations between physical multimorbidity clusters and subsequent depression: cluster and survival analysis of UK Biobank data"

### Figure 1

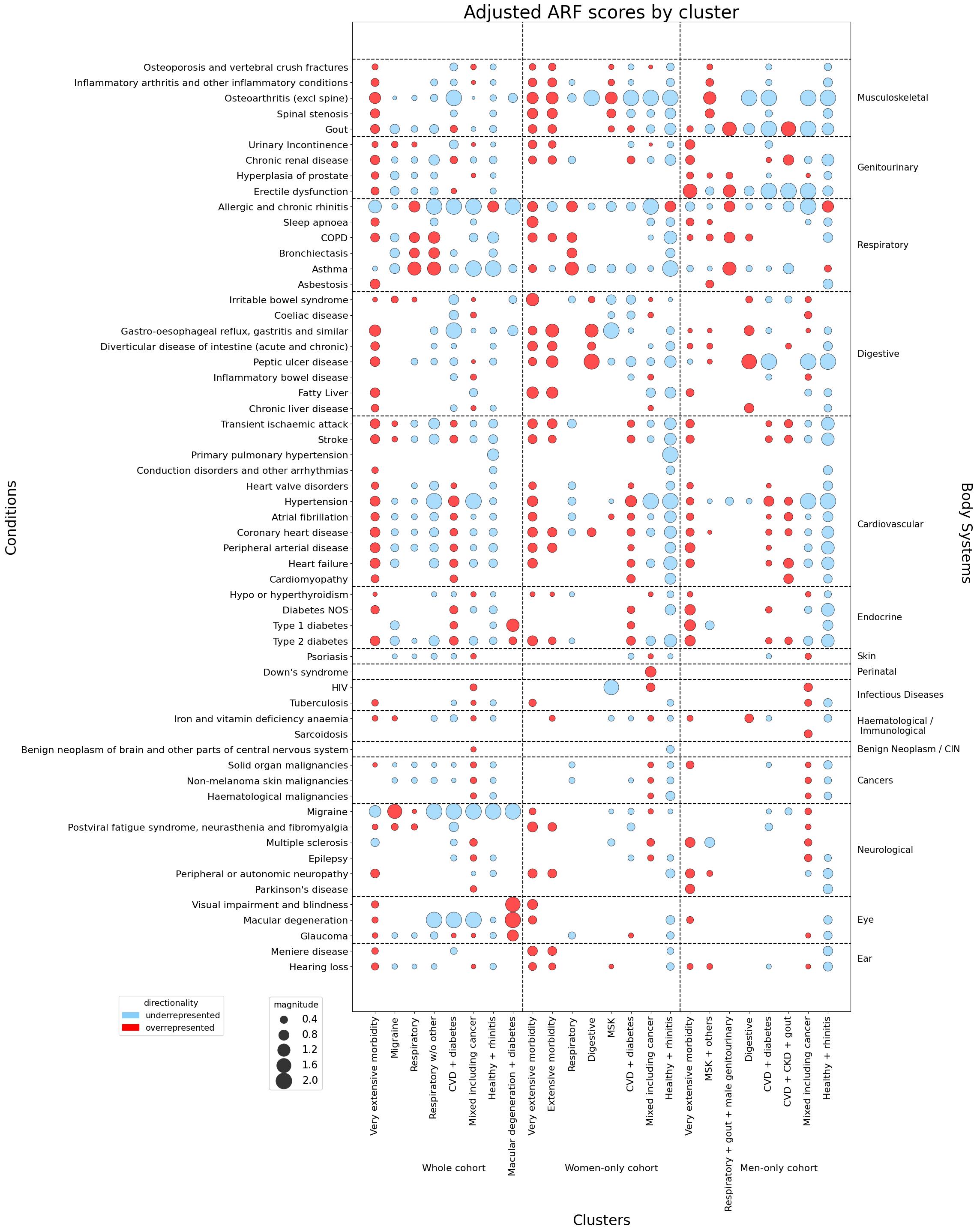

### Figure 2

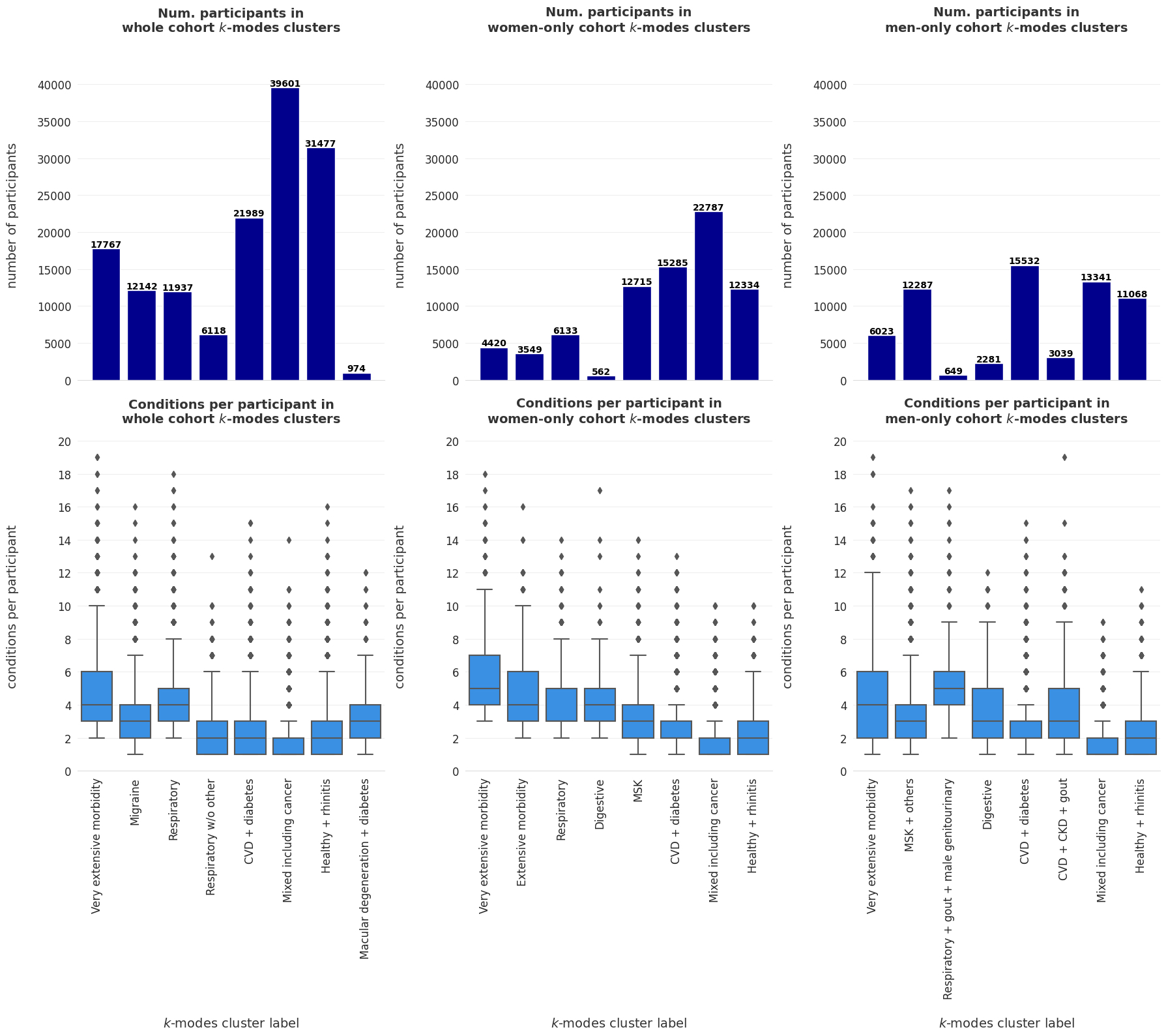

### Figure 3

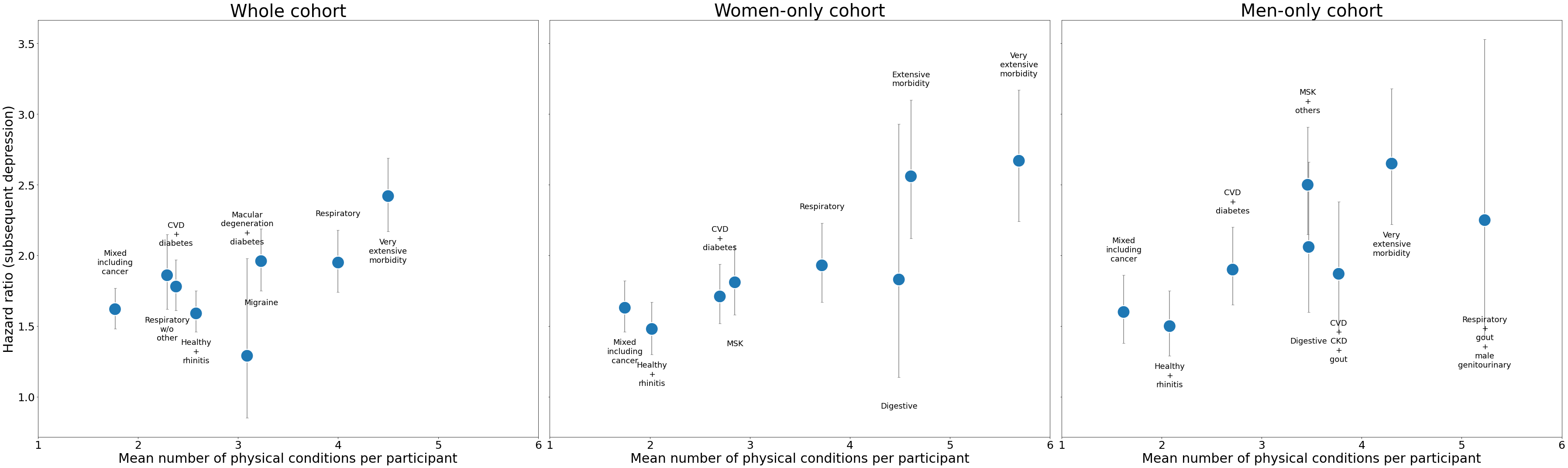

### Supplemental Figure 1

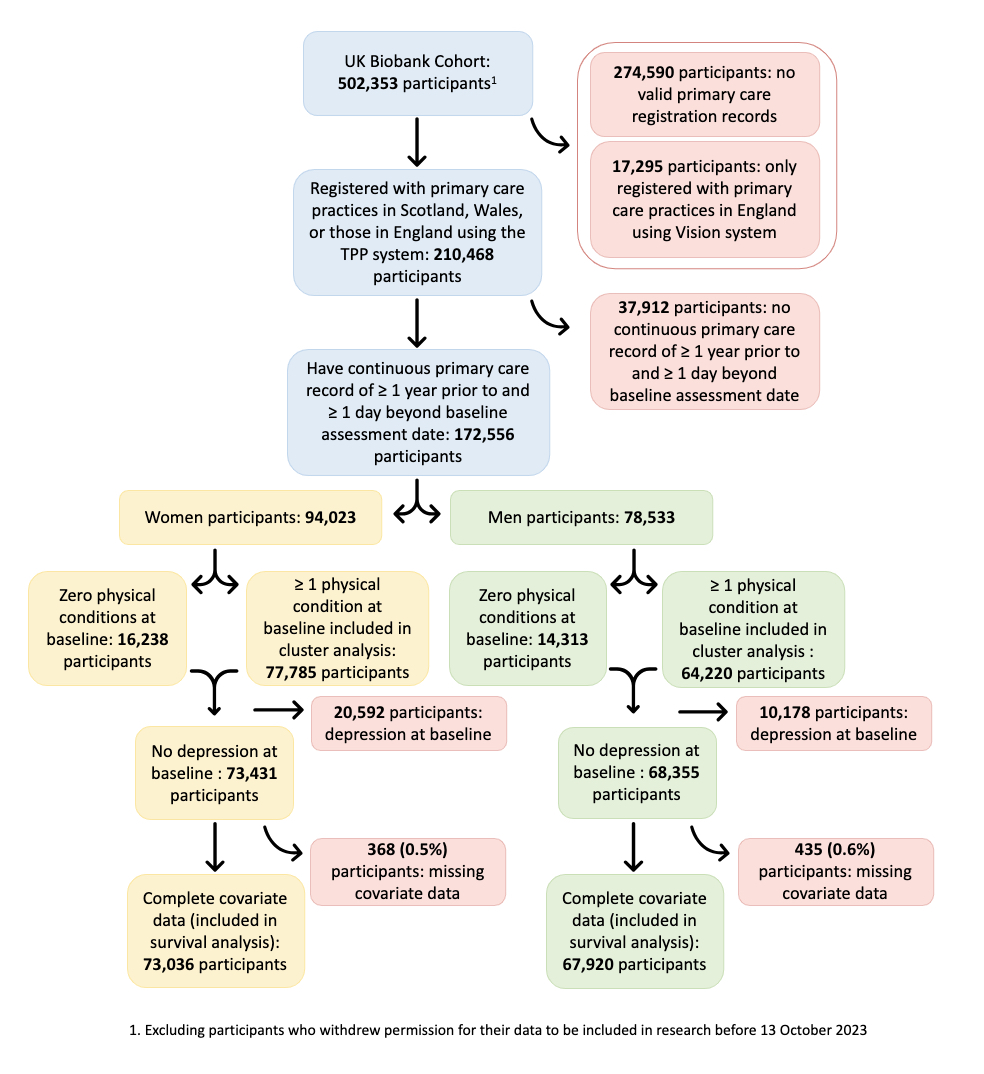

### Supplemental Figure 2

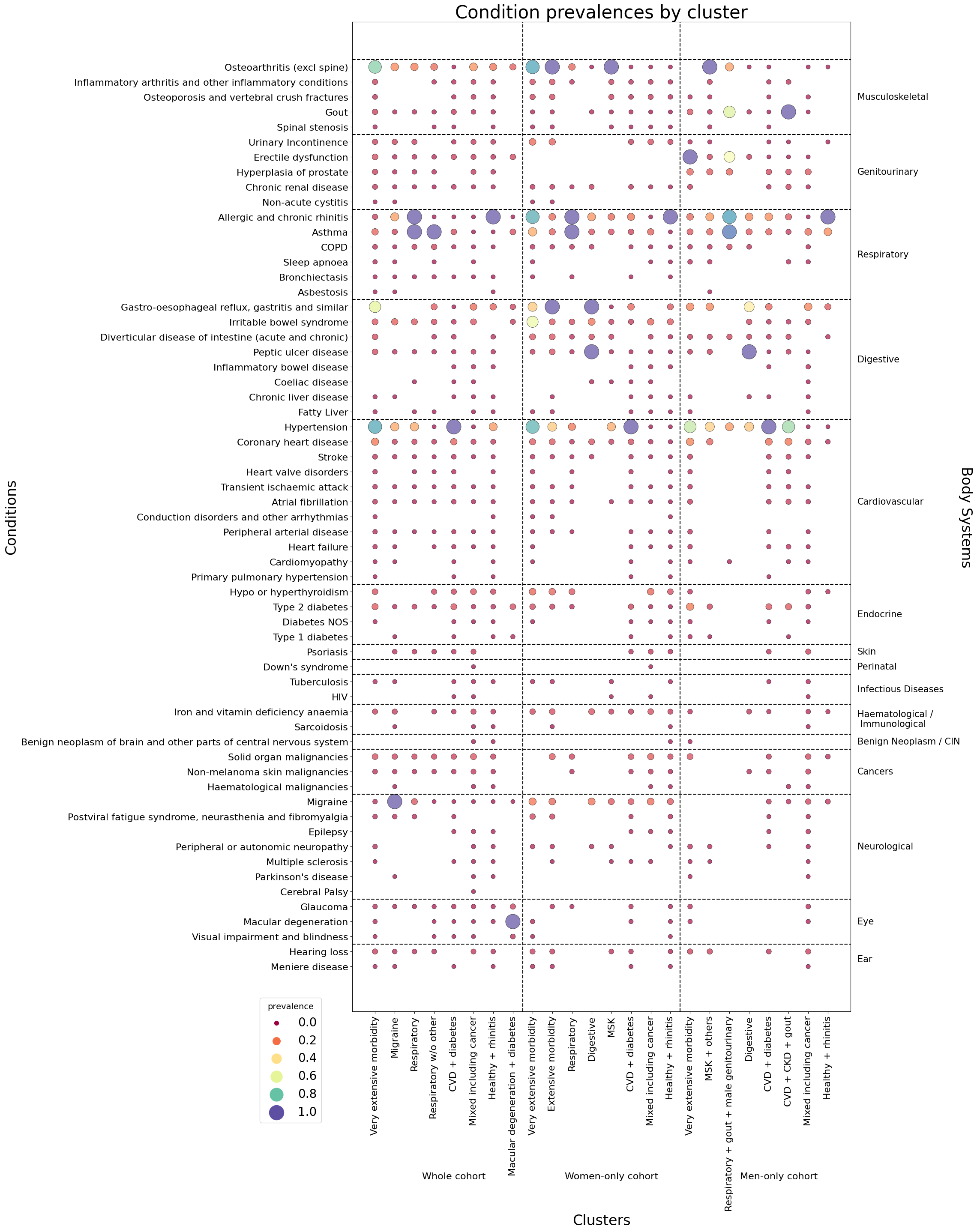

### Supplemental Figure 3

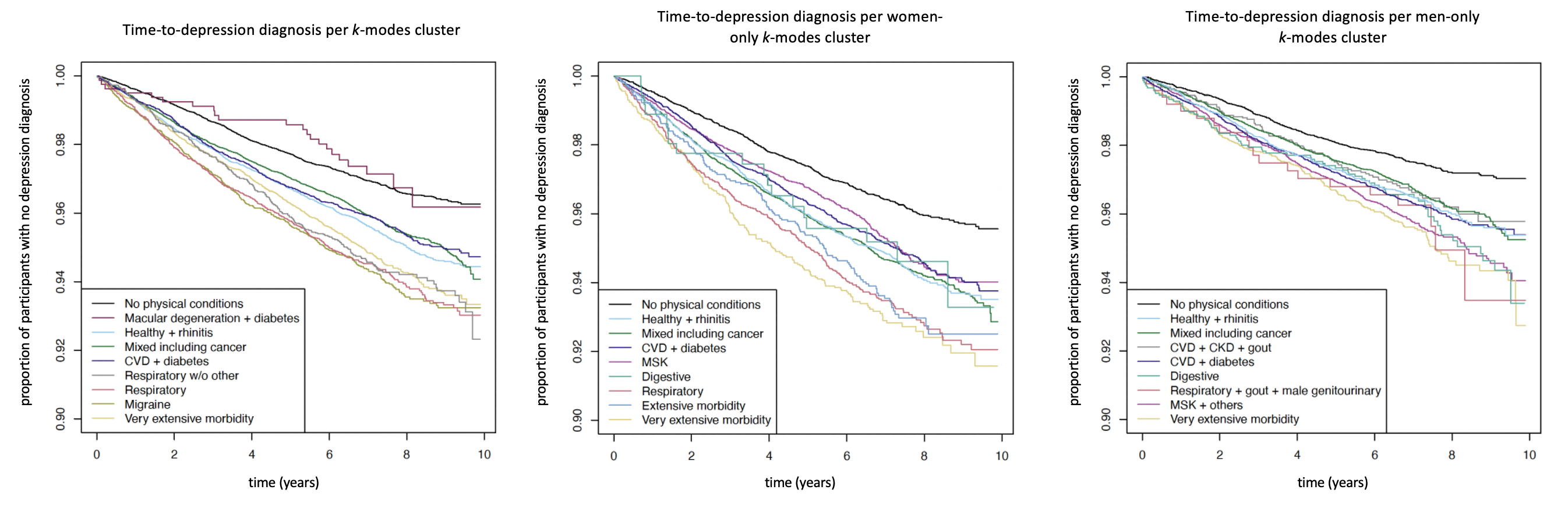
